# Microbial Ecological Signatures Predict Pathogen Emergence and Multidrug Resistance in Cystic Fibrosis Airways up to a Year in Advance

**DOI:** 10.64898/2025.12.28.25342520

**Authors:** Thomas R. Goddard, Jessica AP. Carlson-Jones, Judith Morton, Chee Y. Ooi, Andrew Tai, Morgyn S. Warner, John Wong, Ieuan ES. Evans, Emily Hopkins, Jonathan R. Iredell, Hubertus PA. Jersmann, Katrine L. Whiteson, George Bouras, Michael P. Doane, Nicholas W. Falk, Renee Green, Susanna R. Grigson, Vijini Mallawaarachchi, Belinda Martin, Michael J. Roach, Feargal J. Ryan, Anita Tarasenko, Bhavya Papudeshi, Barbara Drigo, Sarah K. Giles, Clarice M. Harker, Ryan D. Hesse, Riley J. Hodgson, Art Hussnain, Abbey Hutton, Laura K. Inglis, Christopher Keneally, Emma N. Kerr, Craig Liddicoat, Shawn D. Peddle, Carl D. Watson, Qi Yang, Przemysław Decewicz, Peter G. Speck, James G. Mitchell, Elizabeth A. Dinsdale, Robert A. Edwards

## Abstract

Chronic infections in cystic fibrosis (CF) emerge from gradual ecological transitions in the airway microbiome, yet early predictive markers remain poorly defined. We developed a new autoencoder-based framework that outperforms read-based or metagenome-assembled genome-based analyses at capturing the continuum from health-associated commensals to pathogen-dominated, antibiotic-tolerant communities. This improvement is achieved by integrating taxonomic and functional data from 127 sputum and bronchoalveolar lavage metagenomes from 64 people with CF into latent “Clusters of Phylogeny and Functions” (COPFs). Coupled with gradient-boosted random forests, COPFs predicted *Pseudomonas aeruginosa* colonisation, multidrug resistance, and impending infection up to a year before clinical detection. The multidrug-resistant *P. aeruginosa* signature showed the same resistance-mechanism evolution as found in laboratory experiments. The inclusion of eukaryotic markers revealed persistent *Aspergillus fumigatus* signatures even during culture-negative intervals. Applying our South Australian-trained model to over 1,000 global metagenomes from 22 independent CF datasets, we achieved 94% accuracy in predicting *P. aeruginosa* status across platforms and geographies, validating the model’s universal utility. Our results demonstrate that combining datasets with deep learning reveals conserved ecological and metabolic mechanisms in disease progression, transforming metagenomics into a predictive framework for managing chronic infections.

## Introduction

Cystic fibrosis (CF) is a life-limiting genetic disorder caused by mutations in the *CFTR* gene that disrupt chloride ion transport, impair mucociliary clearance, and predispose the lungs to persistent microbial colonisation. Chronic lung infections are the major driver of morbidity and mortality in people with CF (pwCF), despite substantial advances in care and antimicrobial therapy. The introduction of CFTR modulators has transformed clinical outcomes by improving lung function, reducing exacerbations and shifting the airway environment towards one more closely resembling that of individuals without CF^1^. Yet modulators do not fully normalise airway physiology nor eradicate established pathogens, and many pwCF continue to experience recurrent or chronic infections that compromise long-term health^2^.

A central clinical uncertainty is identifying which individuals will progress to disease and when they will have exacerbations and acquire multidrug-resistant (MDR) infections, hampering timely management. Culture-based diagnostics are inherently limited in their ability to detect low-abundance, slow-growing, or emerging pathogens, characterise microbial community dynamics, or capture functional markers such as antimicrobial resistance genes or virulence factors. As a result, no current approach can determine who is at risk or when clinical decline will occur. Disease progression in CF is not instantaneous but rather reflects gradual shifts in the composition and function of the airway microbiome^3^. These ecological transitions include changes in the balance between commensals and pathogens, the emergence of resistant subpopulations, and alterations in metabolic potential, collectively shaping the trajectory of infection^3^.

Here, we analyse thousands of taxonomic and functional features across 127 samples from pwCF. Classical feature selection methods become unstable, and so we employed an autoencoder-based dimensionality reduction approach that preserves co-correlated biological signals while preventing overfitting. Our novel neural network-based framework could identify predictive markers of infection and disease progression from 127 sputum and bronchoalveolar lavage (BAL) metagenomes from our South Australian cohort of pwCF. Our new approach integrates taxonomic and functional analyses into clusters of related features, called COPFs, that we could use to follow microbialization. We trained deep learning models to identify *P. aeruginosa* culture-positive samples and samples containing multidrug-resistant *P. aeruginosa*. We found that resistance mechanisms previously identified *in vitro* are also detectable *in homine*. Uniquely, we determined which individuals were likely to acquire *P. aeruginosa* within the following year. Beyond *P. aeruginosa*, antibiotic exposure left reproducible, class-specific microbial and functional imprints, and the same framework captured fungal and mycobacterial signals that are often missed by culturing. By quantifying medication burden and virulence potential, we defined alternate microbial “health phases” and showed that individuals follow distinct ecological trajectories through this multidimensional space. Finally, we validated our predictions across more than 1,000 metagenomes from 22 global datasets and confirmed that these relationships are robust across populations, genotypes, clinical treatments, sampling sites, environments, and diets. Our analyses demonstrate that conserved microbial and functional signatures precede culture positivity, multidrug resistance, and clinical detection, providing a foundation for predictive diagnostics and personalised intervention strategies to better manage or prevent clinical decline in CF.

## Results

### South Australian cystic fibrosis sampling

South Australia has the highest incidence of CF of any state in Australia, at 21.8 cases per 100,000 people, compared with approximately 11 per 100,000 in the United States^4^ and 16 per 100,000 in the United Kingdom^5^. In 2023 (the most recent year for which data is available), the CF service in South Australia provided care for 388 people with CF.

Between September 2017 and March 2019, we collected sputum (and a single bronchoalveolar lavage (BAL)) samples from 64 different people (16% of the people in the state living with CF), including 35 females (55%) and 29 males (45%). In South Australia, CF care is stratified by age: the Women’s and Children’s Hospital, Adelaide, manages all pwCF under 18 years, after which pwCF transition to the Royal Adelaide Hospital, Adelaide (Table 1). Clinical accessibility and sampling logistics skewed our cohort toward younger individuals, as one of our clinical aspirations is to follow and understand infection progression. Younger pwCF typically experience fewer severe infections and are less frequently colonised by potentially pathogenic bacteria than their older counterparts^6^. The age-based separation is reflected in their lower medication burden, reduced exacerbation frequency, and comparatively simple microbial communities. Because sampling predated the approval of most CFTR modulators in Australia, only one of the participants (pwCF ID 1862551) received modulator therapy (ivacaftor) at the time of sampling (ivacaftor and orkambi had been approved but were only available for a small subset of the pwCF population).

**Table 1.** Clinical and demographic characteristics of the study cohort.

| Characteristic | Value |
| --- | --- |
| Number of participants | 64 |
| Number of samples | 127 |
| Study period | September 2017 to March 2019 (528 days) |
| Sex, female/male | 35 (55%) / 29 (45%) |
| Median age at first sampling (IQR) | 16.5 years (11.0–26.5) |
| Participants <18 years / ≥18 years | 37 (58%) / 27 (42%) |
| Pancreatic insufficiency | 59 (92%) |
| CFRD | 9 (14%) |
| Impaired glucose tolerance | 7 (11%) |
| *P. aeruginosa* infection | 28 (44%) |
| Inpatient samples | 66 |
| Participants with $\geq 2$ samples | 29 |
| CF Genotypes | F508del / F508del: 41 (64%)<br>Unknown / F508del: 7 (11%)<br>F508del / G551D: 4 (6%)<br>Others (1 pwCF each): 12 |

We extracted DNA from sputum and BAL samples and sequenced the DNA of 127 samples using 300 bp MGI paired-end short-read technology, 61 samples from 43 different pwCF were sequenced using the Oxford Nanopore MinION technology, and 10 samples from 9 different pwCF were sequenced using the Oxford Nanopore PromethION technology. We used different sequencing technologies to ensure that our generalised classifier covers current and future sequencing platforms. All the sequences were processed through a similar pipeline to identify the taxonomic profile and functional genes in the sample^7^. We found that each technology detected similar components of the microbial communities (Supplementary Fig. 1). As expected, short-read technology could detect less abundant species in the samples by virtue of increased read depth. In contrast, long-read technology provided more confident functional or taxonomic assignments. We also assembled and annotated 3,215 assembly bins (Supplementary Fig. 2; for this, we included any assembled bins longer than 20 kbp), including 15 metagenome-assembled genome bins that were 90% or more complete, 10% or less redundant, and more than 1,000,000 bp long, and included multiple genomes from the Actinomycetales, Bacteroidales, Bifidobacteriales, Clostridiales, Lactobacillales, and Neisseriales orders.

To distil the highly dimensional signals from bacterial, viral, archaeal, and eukaryotic sources as well as from metagenome-assembled genomes and functional pathways, we applied an autoencoder to reduce a combined dataset (Methods; Tables S1 and S2) into a 50-dimensional latent representation. We correlated the normalised read counts for each taxonomic and functional category with the latent dimensions and used hierarchical clustering on these correlation profiles to identify sets of features with shared covariance structure, yielding biologically interpretable representations of community change. These clusters capture the dominant axes of microbial and functional variation embedded within the latent space, allowing us to treat each as a single meta-feature for downstream analyses. We refer to these integrated sets as Clusters of Phylogeny and Functions (COPFs) (see Methods). Details of autoencoder training and feature clustering are described in *Methods: Data reduction* and *Gradient-boosted Random Forest*, and the code is available online^8^.

### Microbial Community Shifts Along a Treatment–Health Gradient

Our metadata captures multiple indicators of overall health in pwCF, including their lung function (as measured by forced expiratory volume in 1 second (FEV_1_) at the time of sampling, and the proportion of that FEV₁ compared to their best FEV₁ from the previous 12 months, together with the number and classes of antibiotics, antifungals, steroids, and other medications prescribed, and the interval since the last hospitalisation. Antibiotic administration was recorded for 82 of 127 samples (65%). Inhaled tobramycin was the most frequently prescribed in our CF cohort. Oral therapies were dominated by itraconazole, amoxicillin-clavulanate, and ciprofloxacin, whereas during acute exacerbations, piperacillin-tazobactam and tobramycin were predominantly prescribed as intravenous and/or inhaled therapies. We found that COPFs, driven by *Achromobacter* and other *Alcaligenaceae*, were consistently enriched in antibiotic-exposed samples, whereas COPFs containing *Gemella*, *Pasteurella*, and *Neisseria* were characteristic of antibiotic-free samples. Functional signatures of antibiotic exposure in our COPFs included ectoine and hydroxyectoine catabolism, consistent with osmoprotectant metabolism under β-lactam and clavulanate therapy. However, our data includes more than just antibiotics and antifungals. To integrate all drug prescription measures, we developed a unified principal-component framework that combined antibiotic, antifungal, steroid, and monoclonal antibody use (omalizumab (anti-IgE) for ABPA when steroids were not tolerated) with the FEV₁ ratio, generating a single *Medications Score* that reflects the aggregate treatment burden at the time of sampling (Fig. 1a). This multidimensional index correlates strongly with the number of prescribed therapeutics, but correlates with neither lung-function measures nor inpatient status (Supplementary Fig. 3). We hypothesise that, in clinical practice, treatment intensity and lung function are only loosely coupled, as pwCF under active management may be recovering from an exacerbation and temporarily exhibit improved respiratory performance compared with those not currently undergoing treatment. Moreover, numerous independent factors influence FEV_1_ fluctuations, including extrapulmonary manifestations such as CF-related diabetes and nutritional status.

**Fig. 1.**
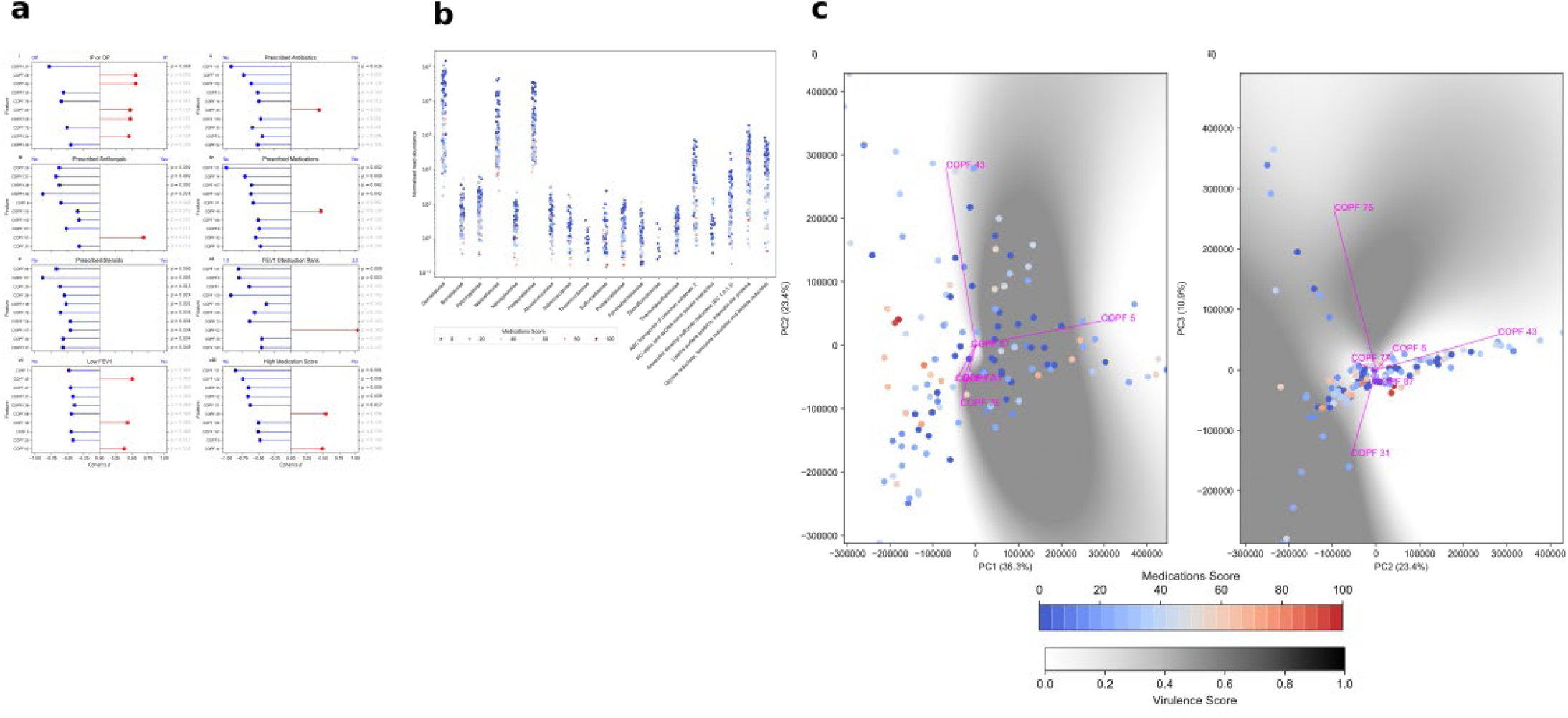
The Medications Score shows an infection continuum. **a.** Comparison of the top ten COPFs that discriminate health measures, including the Medications Score. For each category the directionality is shown on top of the bar plot. The *x*-axis has the effect size (Cohen’s *d*) for each comparison, and the *y*-axis has the COPFs with the biggest effect for each measure. The T-test *p* value adjusted for multiple testing using the Benjamini-Hochberg procedure is shown on the right, and adjusted *p*-values < 0.05 are highlighted. (*n* = 125) **b.** The normalised read abundance for each of the functions and phylogenies in COPF 131. The abundance is coloured by medication score from blue (least) to red (highest medication score) (*n* = 125). **c.** Comparison of COPF distributions to virulence metrics. PCA of the COPF distribution showing the relationship between principal components 1 and 2 (i) and 2 and 3 (ii). The points are coloured by the medication score, with blue indicating fewer and red indicating more prescribed medicines. The virulence metric shades the mesh interpolated between the COPFs, with green indicating fewer and brown indicating more virulence-associated terms. (*n* = 125).

PERMANOVA revealed a significant difference among the 23 groups of Medications Score (pseudo-*F* = 1.43, *P* = 0.036, n = 125, 999 permutations; Supplementary Fig. 4), so we compared effect size (Cohen’s *d*) for each COPF for the various health-related metrics using dichotomous data groups (Fig. 1a). A consistent subset of COPFs shows significant differences across multiple health-related measures. For example, sequences mapping to COPF 131 are more abundant in outpatients than inpatients; when individuals are not on antibiotics, antifungals, or other medications; and when individuals have a lower FEV_1_ obstruction rank. This cluster contains normal oral flora, including Neisseriaceae and Gemellaceae (Fig. 1b). Other clusters associated with health (Supplementary Fig. 5) also lack common pathogens and virulence phenotypes.

We hypothesised that variation in medication metrics would be reflected in corresponding differences in microbiome virulence potential. To evaluate this, we devised a quantitative measure of virulence and applied it to each COPF. Specifically, we calculated a Jaccard Index between the taxonomic and functional annotations identified in each COPF and those catalogued in the Virulence Factor Database^9^. The resulting index was scaled from 0 (fewest virulence-associated annotations) to 1 (highest representation of virulence-associated annotations) and subsequently projected onto principal component analysis (PCA) plots (Fig. 1c). Due to inherent limitations in visualising this multidimensional metric within two dimensions, we present the first three principal components, which together explain 70.6% of the total variance.

Comparison of each COPF’s gene content against the Virulence Factor Database showed a progressive enrichment of virulence-linked functions, including secretion systems, iron acquisition, and stress-response modules, as treatment burden increased. No single microbiome configuration defined a healthy or diseased configuration; rather, pwCF communities occupied a continuum of partially overlapping phases with differing virulence potential. This gradient reflects the ecological restructuring that accompanies repeated therapy and progressive airway decline, as well as the wide interindividual heterogeneity^10^.

Longitudinal sampling of six pwCF revealed highly idiosyncratic temporal dynamics at the taxonomic level (Fig. 2a). Across the cohort, we observed a consistent ecological trajectory as lung function deteriorated, reminiscent of disturbance-driven collapse in macro-ecosystems (Fig. 2b). However, using the medications metrics score (Fig. 2c), each individual followed a unique path through compositional space, alternating between transiently stable and unstable phases until the next perturbation. None converged toward a common community state. Recurrent insults such as antibiotic exposure, inflammation, and structural damage, destabilise resident commensals, including *Neisseria*, *Gemella*, and *Prevotella*, which are gradually replaced by opportunistic pathogens, including *P. aeruginosa*, *Staphylococcus aureus*, and *Streptococcus* spp. This transition unfolds through intermediate, unstable states that mix remnant commensals with encroaching pathogens shaped by individual treatment histories. We hypothesise that opportunists rapidly colonise disturbed mucosa and outcompete health-associated taxa, but these shifts remain subtle and heterogeneous, making them difficult to detect in individuals.

**Fig. 2.**
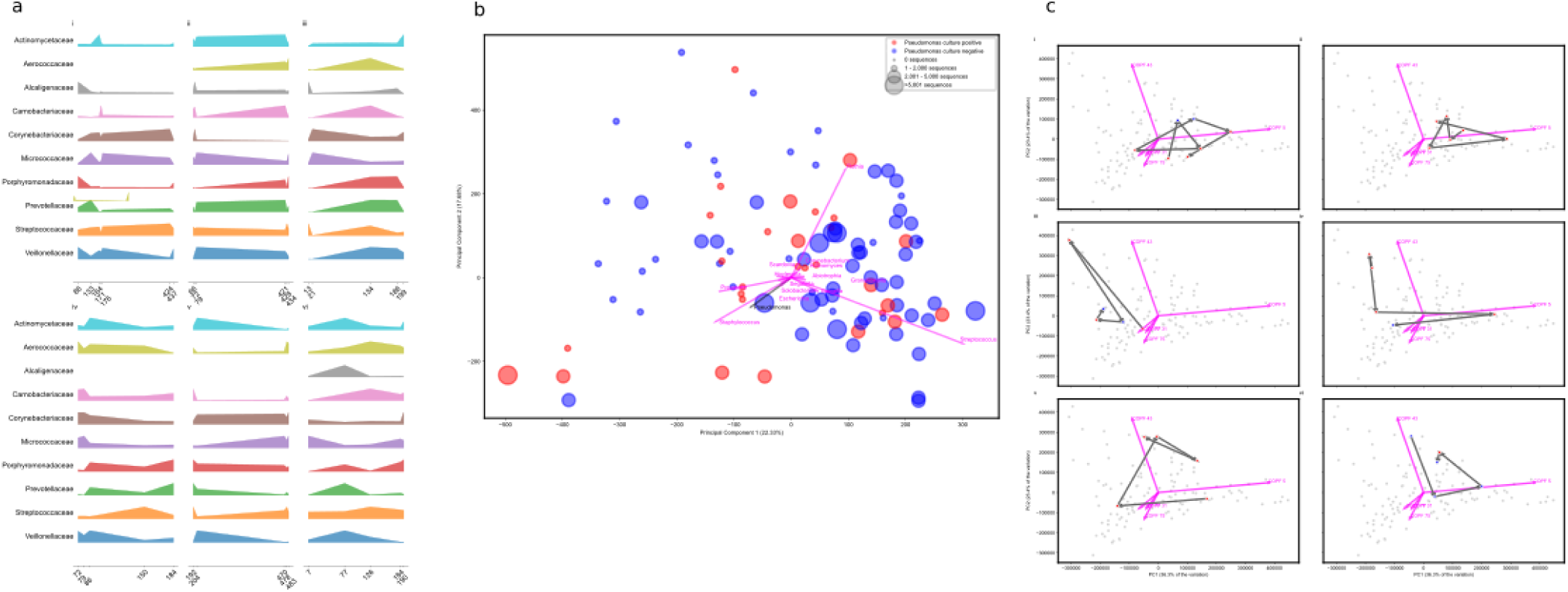
The microbiome of every person is different. **a.** Ridge plots of the microbiome of different people sampled over time, showing the abundance of the top 10 microbial taxa over the days since the first sample in the project was collected; pwCF IDs i) 788707; ii) 825012; iii) 676138; iv) 785991; v) 748699; vi) 658355 **b.** PCA of microbial taxonomy. For each point the size of the dot reflects the normalised number of reads that map to *Pseudomonas* while the colour represents the *P. aeruginosa* culture status. The median number of sequences that mapped to *Pseudomonas* was 2,721 per sample. The top 15 PCA taxonomic loadings are shown to identify which taxa drive the separation of the microbial communities across these axes (*n* = 127). **c.** Progression of phase shifts in microbial communities. The same PCA ordination of all COPFs is shown for each pwCF. For each pwCF, highlighted points are coloured by whether they were seen as an out-patient (blue) or in-patient (red) in each panel. The arrows indicate the sampling progression in time. The x (PC1) axis explains 36.3% of the variation and the y (PC2) axis explains 23.4% of the variation. The pwCF IDs are in the same order as panel a. (*n*=125)

### Predicting *P. aeruginosa* presence from latent microbiome features

Next, we sought to assess whether our model could predict *P. aeruginosa* culture status. In our cohort, *P. aeruginosa* was cultured from 36 samples from 28 individuals, whereas it was not cultured from 91 samples. Eight individuals had at least one sample where *P. aeruginosa* was not cultured, but others in which it was.

Among the most discriminatory predictors of *P. aeruginosa* culture status is COPF31 (Supplementary Table 3), which contained the Pseudomonadaceae family. However, this importance was not driven solely by the taxonomic signals, as the cluster contains several functional subsystems, including the Zot toxin, phenazine biosynthesis, and pyoverdine synthesis. Zot increases the permeability of epithelial cells by reversibly disrupting tight junctions^11^. The phenazines are secondary metabolites, including pyocyanin, that act as antibiotics, virulence factors, and survival factors, and may serve as electron acceptors in the CF lung^12,13^. Pyoverdine is an iron-scavenging siderophore essential for *Pseudomonas* colonisation of the CF lung^14^. Thus, while directly detecting *Pseudomonas* sequences strongly contributes to determining whether *P. aeruginosa* would be cultured from a sample, functional features and other taxonomically diverse signals also proved informative.

COPF 132 (Supplementary Table 4) is also a strong predictor of *P. aeruginosa* culture status (Fig. 3). The key signals in this cluster include cyanate hydrolysis, which *Pseudomonas* uses to outcompete *Staphylococcus*^15^, glycerolipid and glycerophospholipid biosynthesis which *Pseudomonas* (and other bacteria) use to overcome antibiotic treatments^16^, one-carbon metabolism by tetrahydropterines that create the cofactor for phenylalanine hydroxylase in *P. aeruginosa*^17^, and the HigAB toxin-antitoxin subsystem, the products of which regulate the production of the virulence factors pyochelin and pyocyanin, and also regulate swarming and biofilm formation in *P. aeruginosa*^18^. The abundance of each of these subsystems across the different samples is correlated, and they are equally discriminatory.

**Fig. 3.**
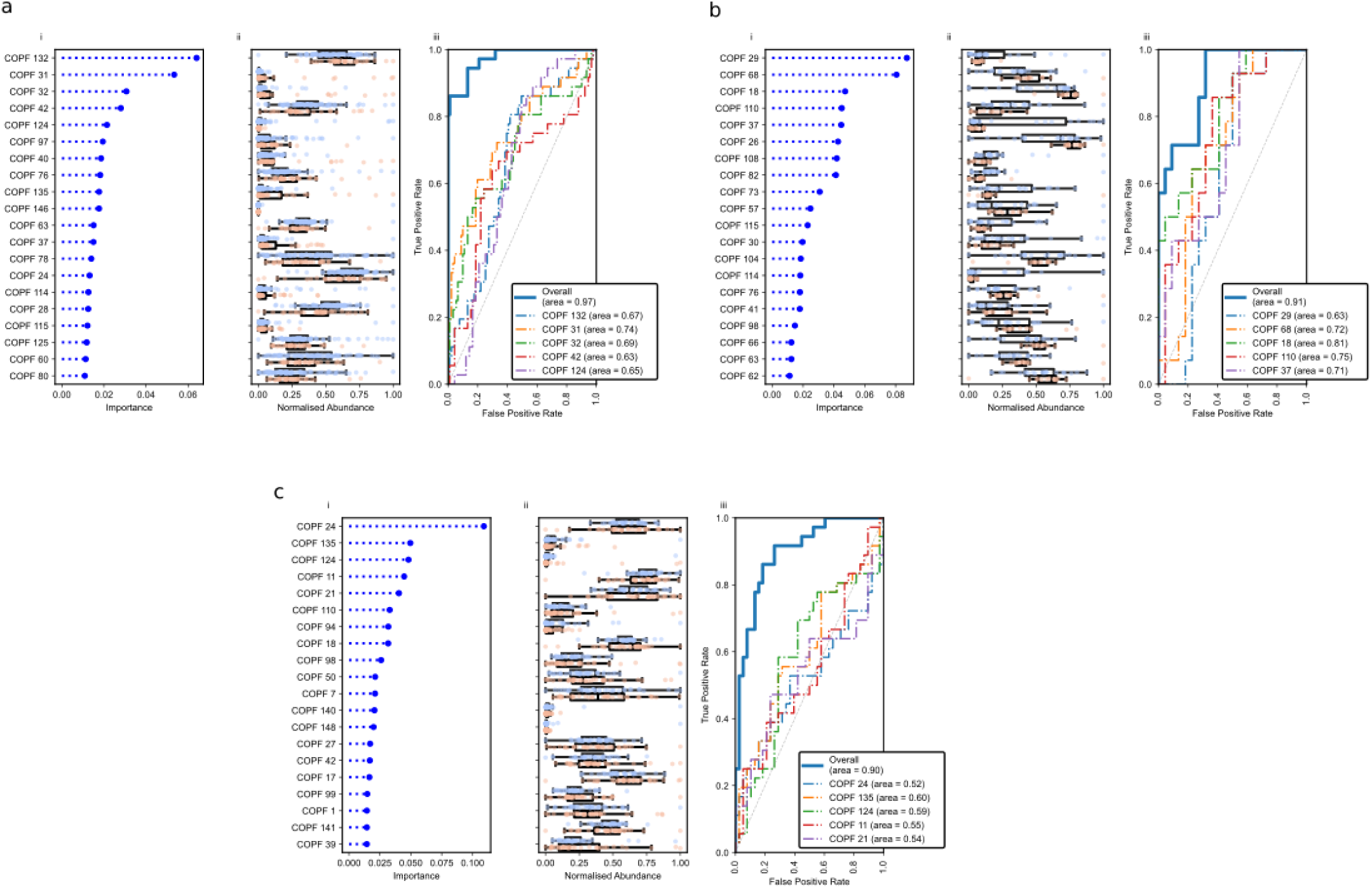
Predicting clinical outcomes from metagenomics data. **a.** the prediction of *P. aeruginosa* culture status (*n* = 127), **b.** the prediction of multi-drug resistant *P. aeruginosa* (*n* = 36), and **c.** the prediction of acquisition of *P. aeruginosa* in the next twelve months (*n* = 74). Each triptych contains (i) the gradient-boosted random forest feature importance plot for each COPF, (ii) the abundance of those COPF in samples with (red) or without (blue) the phenotype, and (iii) the ROC curve for the top 5 COPFs at predicting the phenotype and the overall ROC curve for all data. Area under the curves (AUC) are included in the ROC legend.

Receiver operating characteristic (ROC) curves for the five most informative clusters and the overall predictive power of the model (Fig. 3a) support that the prediction of *P. aeruginosa* culture status is not explained by *Pseudomonas* sequences alone, but by a combination of taxonomic and functional features that collectively enhance predictive accuracy. When trained on all COPFs, the classifier achieved an AUC of 0.97, demonstrating that the predictive signal for *P. aeruginosa* culture status is deeply embedded in the data.

Among the 22 MAGs annotated as *Pseudomonas* by at least one taxonomic classifier, 14 had statistically significant abundance differences and were enriched in the culture-positive samples when comparing *P. aeruginosa* culture-positive samples versus culture-negative samples (Fig. 4).

**Fig. 4.**
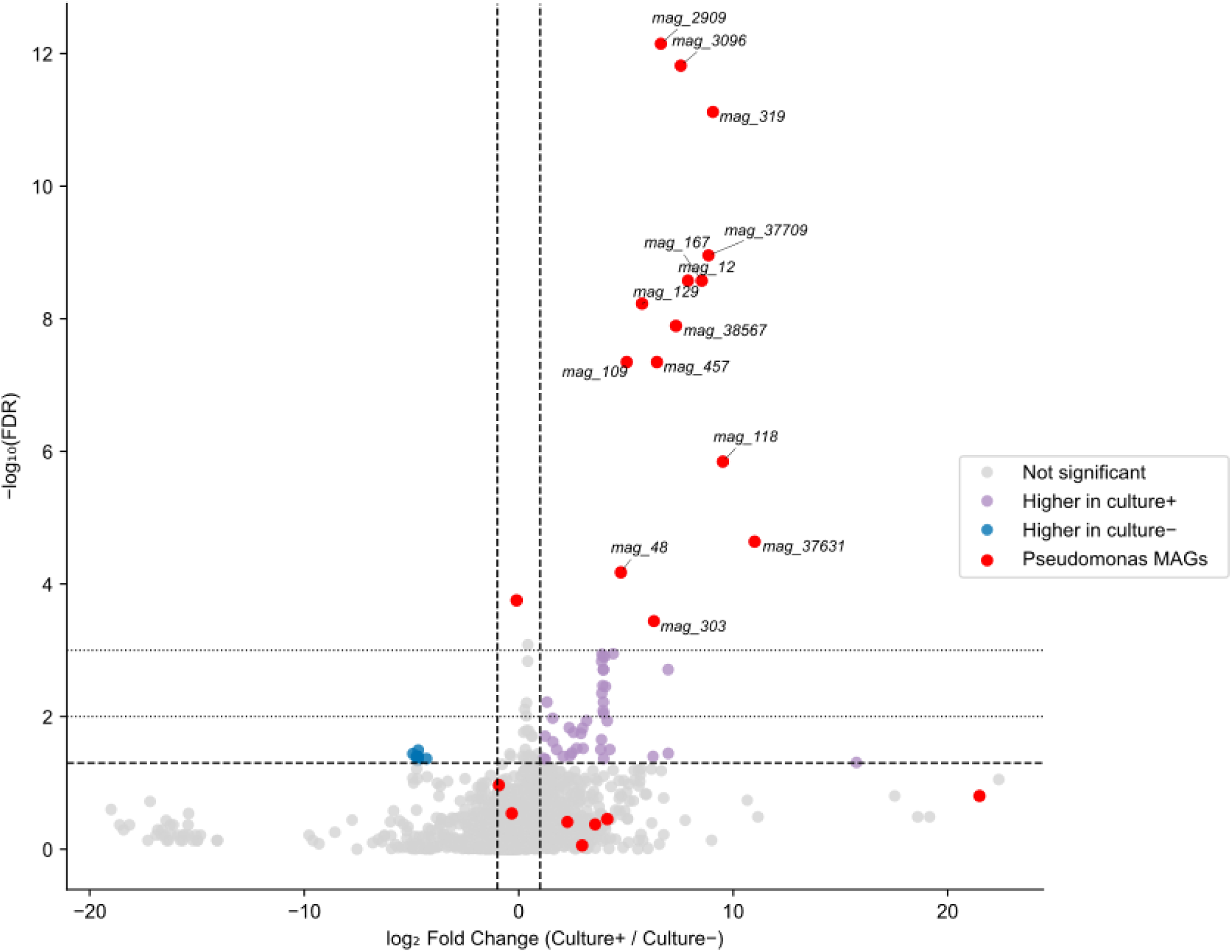
Identification of assembly bins that reflect *P. aeruginosa* culture status. A volcano plot of all (*n* = 3,215) metagenome assembly bins showing the log₂ fold change in median RPKM between samples that were culture positive and negative for *P. aeruginosa*. Each point represents a single bin, with the x-axis indicating the direction and magnitude of differential abundance and the y-axis showing the −log₁₀ false discovery rate (FDR) from a two-sided Mann–Whitney U test corrected for multiple comparisons. Horizontal dashed and dotted lines mark FDR thresholds of 0.05, 0.01, and 0.001, respectively. Vertical dashed lines indicate a twofold change in abundance (|log₂FC| = 1). Bins significantly enriched in *P. aeruginosa*-positive samples are shown in violet, those enriched in *P. aeruginosa*-negative samples in green, and non-significant bins in grey. MAGs annotated as *Pseudomonas* by at least one taxonomic approach are highlighted shown in red. Bins significantly enriched in *P. aeruginosa*-positive samples and annotated as *Pseudomonas* are labelled.

**Fig. 5.**
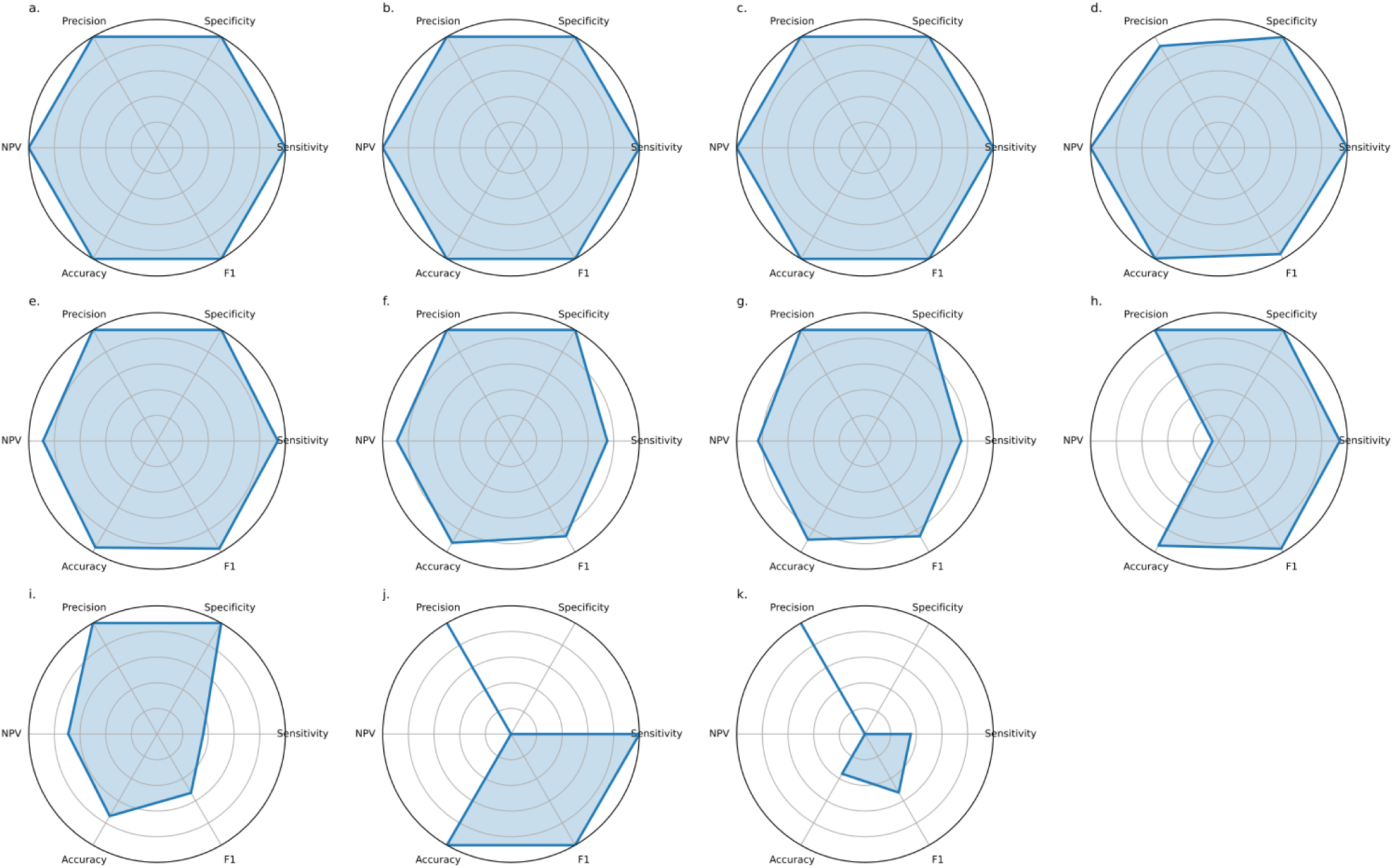
Performance profiles for eleven independent CF BioProjects. summarised as multimetric radial plots, with each panel representing sensitivity, specificity, precision (PPV), negative predictive value (NPV), accuracy, and F_1_ score scaled between 0 and 1. Each polygon depicts the classification performance for an individual cohort based on its true-positive (TP), true-negative (TN), false-positive (FP), and false-negative (FN) counts. a. PRJNA516442 (*n* = 93); b. PRJNA316588 (*n* = 18); c. PRJNA1126024 (*n* = 2); d. PRJNA931830 (*n* = 260); e. PRJEB32062 (*n* = 27); f. PRJNA644285 (*n* = 12); g. PRJEB51171 (*n* = 64); h. PRJNA1081394 (*n* = 549); i. PRJNA1055940 (*n* = 61); j. PRJEB14440 (*n* = 5); k. PRJNA510441 (*n* = 14)

Although the full genome size of *Pseudomonas* is typically 5.5–7 Mb^19^, the assembled fragments we recovered ranged from ∼25 kb to a whole MAG of ∼6.5 Mb, with a median assembly size of ∼42 kb, suggesting considerable genome fragmentation.

### Microbiome features associated with multidrug-resistant *P. aeruginosa*

Having established that our models can robustly predict *P. aeruginosa* colonisation, we next asked whether the same framework could discriminate between multidrug-resistant (MDR) and non-MDR isolates. We classified *P. aeruginosa* as MDR if it was either reported as MDR by SA Pathology or was resistant to two or more classes of antipseudomonal agents. Among the 36 samples that were culture-positive for *P. aeruginosa*, 14 harboured MDR isolates. To investigate the microbial and functional features associated with MDR phenotypes, we compared these 14 samples with 22 culture-positive, non-MDR cases (Fig. 3b).

COPF 82 was among the clusters that distinguished MDR from non-MDR samples. That cluster only contains two subsystems: “NADH ubiquinone oxidoreductase vs. multi-subunit cation antiporter” and “Menaquinone biosynthesis from chorismate via 1,4-dihydroxy-6-naphthoate”. The first subsystem contains the 14 genes that encode the different subunits of the NADH ubiquinone oxidoreductase (EC 1.6.5.3), the seven genes that encode subunits A-G of the Na(+) H(+) antiporter, and the multisubunit-sodium/proton-antiporter-like complex, protein X. *P. aeruginosa* carries three NADH:quinone oxidoreductases (NQR, NUO, and NDH2), and experimental deletions of these enzymes mimic the transition from exponential growth to stationary phase, driving increased biofilm formation and pyocyanin production, hallmarks of heightened virulence. Experimentally, deletion mutants exhibit enhanced lethality in macrophage and murine infection models^20^. Mutations in NADH ubiquinone oxidoreductase (*nuo*) genes increase resistance to antibiotics, including tobramycin^21^, and have repeatedly emerged in CF isolates^22^. Thus, the abundance patterns of this COPF in our metagenomes mirror the experimental evidence, suggesting that respiratory chain remodelling is under active selection in South Australian pwCF, potentially facilitating the emergence of multidrug-resistant *P. aeruginosa*.

Across these and the other important features distinguishing multidrug-resistant from non-resistant *P. aeruginosa*, a consistent theme emerged: metabolic re-engineering rather than the acquisition of single canonical resistance genes. Several COPFs highlight complementary aspects of this transition. COPF 29, which is less abundant among samples with MDR isolates, includes a β-carbonic anhydrase, lipid metabolism genes, and universal stress proteins: functions that typically maintain pH balance and general stress tolerance. In contrast, COPF 68 and COPF 18 were enriched in MDR samples and encompassed enzymes connecting glycolysis, the pentose-phosphate pathway, and nucleotide biosynthesis, as well as modules for Fe–S cluster assembly, RNA modification, and capsule formation. These collectively point to strengthened redox management, DNA repair, and translational control. The accompanying enhancement of pyrimidine synthesis, sulfosugar metabolism, and biofilm-associated genes implies increased anabolic and structural capacity. Together, these signatures indicate that MDR *Pseudomonas in homine* reinforce central metabolic and informational networks to sustain growth and resilience under antibiotic stress, rather than relying solely on dedicated resistance determinants.

### Predicting pwCF progression to *P. aeruginosa* colonisation

Having defined the microbial features associated with established MDR phenotypes, we next asked whether the same framework could identify signatures that precede colonisation, focusing on pwCF who were initially culture-negative for *P. aeruginosa* but subsequently acquired the pathogen. Our dataset included 36 samples from 18 individuals who were baseline culture-negative for *P. aeruginosa* but would convert to culture-positive within the following 12 months, and we compared them to 38 samples from 20 individuals who remained culture-negative over the same period (Fig. 3c, Supplementary Table 5).

Microbial communities preceding the acquisition of *P. aeruginosa* displayed coordinated shifts in metabolism and cell-envelope physiology, rather than the emergence of a single precursor species. The COPFs most enriched in pwCF who converted to *P. aeruginosa* positivity (COPF 110, 24, 11; Fig. 3c, Supplementary Table 6) each captured different facets of this transition. COPF 110 includes iron(III)-dicitrate transport systems and xanthine dehydrogenase subunits, indicating intensified iron acquisition and envelope remodelling (modifications to the outer membrane and wall architecture), which enhance stress tolerance and promote pathogen establishment. COPF 24 encompasses aromatic- and lysine-biosynthetic pathways, purine metabolism, and expands anabolic and structural capacity for peptidoglycan crosslinking, nucleotide synthesis, and stress signalling. COPF 11 includes fatty-acid and cofactor biosynthesis modules, such as enoyl-ACP reductases and NAD/NADP generation, supporting membrane biogenesis and intrinsic tolerance. Collectively, these features describe a microbial milieu increasingly configured for redox buffering, membrane renewal, and metabolic flexibility, conditions that pathogens routinely exploit to colonise the CF airway. Although individual COPFs provided modest discrimination (area under the curve (AUC) ≈ 0.5–0.6), their aggregate signal achieved an AUC of 0.9 (Fig. 3c), underscoring that conversion to *P. aeruginosa* positivity reflects a community-wide ecological shift toward a pathogen-permissive state rather than a singular microbial harbinger.

### Predicting other pathogens

We tested whether the same analytical framework could be used to predict the presence or emergence of other bacterial pathogens commonly detected in the CF airways. Our cohort contained many younger people (the median age at first sampling was 16.5 years). More aggressive CF treatments have reduced the incidence of infection, and age is a primary indicator of infection severity. Consequently, it was less common to detect CF-associated pathogens, including non-tuberculous *Mycobacterium* (11 positive samples), *Stenotrophomonas maltophilia* (9 positive samples), *Achromobacter xylosoxidans* (3 positive samples), and *Burkholderia* (1 positive sample) in our cohort (Supplementary Table 7).

The non-tuberculous *Mycobacterium* (NTM) detections include six *Mycobacterium abscessus* detections from six individuals, along with three observations of *M. intracellulare* from two individuals, and one example of *M. terrae* (Supplementary Tables 7 and 8). There was one sample described as not speciated with further speciation not possible using 16S. Three out of the eleven NTM-positive samples were from patients just commenced or about to commence NTM eradication therapy. All three were smear-positive, and all met the ATS criteria for NTM pulmonary disease^23^. None of the others met those criteria. These low observation frequencies limit the performance of deep learning models, and consequently, detecting *Mycobacterium* was more challenging than detecting *Pseudomonas* (Supplementary Fig. 6). Nonetheless, several clusters were predictive of *Mycobacterium* culture status, primarily through their absence. In particular, the commensal, facultatively aerobic genera that are characteristic of well-oxygenated mucosal environments, including *Pasteurella, Neisseria, and Gemella,* were inversely associated with *Mycobacterium* presence. As we showed above, these taxa typically dominate less inflamed, antibiotic-naïve communities, in contrast to the redox-stressed, lipid-rich, and biofilm-associated niche that favours non-tuberculous *Mycobacterium*. The underlying antagonism may arise from overlapping ecological pressures, including competition for iron and heme, oxidative and bacteriocin-mediated inhibition by *Neisseria* and *Gemella*, and differential responses to chronic inflammation and antibiotic exposure.

Detection of *S. maltophilia* highlights one of the inherent challenges of clinical metagenomics for pathogen identification (Supplementary Fig. 7). In most samples where *S. maltophilia* was cultured, there was concordance between sequencing and culturing. However, in others, *Stenotrophomonas* sequences were detected only at the threshold of detection. For example, samples from pwCF 785991 and 770560 were dominated by *Streptococcus* sp., with very few sequences indicating *Stenotrophomonas*. In contrast, samples from pwCF 785991 and 1470026 were dominated by *Rothia*, despite culture-based evidence of *S. maltophilia* in both cases. In these cases, the high selectivity of bacterial culturing vastly exceeds the sensitivity of metagenomics.

### Fungal detection

There are multiple challenges in detecting fungal taxa in CF airway samples, reflecting both the biology of fungal colonisation and the confounding effects of antifungal therapy. We examined this complexity in detail for one individual (pwCF 676138; Supplementary Table 9), who had *Aspergillus fumigatus* detected across multiple sampling dates. Although *A. fumigatus* reads were consistently identified across all metagenomes, the organism was only cultured on two occasions. Because *A. fumigatus* was cultured from this pwCF both before and after these specific dates, the negative cultures likely reflect suppression of fungal growth by antifungal therapy rather than clearance of the organism. Given that colonisation is common, even if a single isolated colony of *A. fumigatus* is cultured, it may not be reported as clinically significant. In this instance, and in contrast to *S. maltophilia* detection, metagenomic detection provides a more reliable indicator of fungal persistence than culture status.

We next asked whether metagenomic detection aligned with immunological markers routinely measured in pwCF to identify *A. fumigatus* sensitisation or allergic bronchopulmonary aspergillosis (ABPA). Because there is no definitive diagnostic test for ABPA, we classified our samples using the revised International Society for Human and Animal Mycology (ISHAM)-ABPA working group consensus criteria for diagnosing ABPA^24^ (Methods; Supplementary Table 10). Our cohort included 51 samples from 35 different individuals with specific IgE measurements. 7 samples (14 %) were classified as having ABPA, 26 (51 %) as being sensitised, and 18 (35 %) as not sensitised (Supplementary Table 10).

We incorporated COPFs spanning all phylogenetic markers, including both bacterial and eukaryotic taxa (which we designate COPFe). ROC analyses of the five most predictive clusters yielded only modest individual performance (AUC = 0.52–0.80; Supplementary Fig. 8), indicating that no single taxonomic or functional axis alone captures the ABPA signature. In contrast, when all COPFs were combined, the ensemble model achieved an AUC of 0.98, demonstrating excellent predictive accuracy. Together, these results suggest that antifungal exposure and the patchy distribution of fungal elements within sputum continue to limit diagnostic sensitivity.

### Global validation of predictive metagenomics using publicly available CF datasets

Our work presents an entirely new way to analyse metagenomics sequence data. To demonstrate the robustness of our analysis, we built predictive models using COPFs to identify who is likely to be colonised with *P. aeruginosa*, who has MDR *P. aeruginosa,* and who will be colonised with *P. aeruginosa* in the future. We could only test the detection of *P. aeruginosa* using previously published metagenomic data, as we were unable to identify any studies that included metadata on MDR status or future colonisation by *P. aeruginosa*. We analysed more than 2 terabases of publicly available CF metagenomic data, spanning over 1,500 sequencing runs from 22 BioProjects deposited in the Sequence Read Archive (SRA) (Supplementary Tables 11 and 12; Fig. 5). Datasets were identified through a systematic search of SRA metadata for CF-related metagenomes or metatranscriptomes, excluding amplicon libraries, and processed uniformly using the same atavide-lite pipeline that was used to process our samples from South Australia.

Across all projects, our model correctly predicted *P. aeruginosa* culture status for more than 90 % of the 1,500 samples with available clinical metadata. Accuracy remained high across sequencing platforms (Illumina, BGI, or Oxford Nanopore) and sample sources (sputum, bronchoalveolar lavage, or nasal/oropharyngeal swab), with most misclassifications attributable to datasets with incomplete culture records, very low read counts, or single-end sequencing.

Re-analysis of the sputum metagenomes, which compared CF, COPD, smokers and healthy subjects (PRJNA316588), correctly classified all 17 high-quality runs, reproducing their *P. aeruginosa*-positive and negative assignments (11.8 % culture-positive)^25^, and suggesting that our models may also be appropriate for non-CF bronchiectasis. Similarly, the depletion experiment (PRJNA516442) was perfectly recapitulated: all 15 metagenomes were correctly called, including the loss of the *P. aeruginosa* signal in Molysis-treated samples^26^.

Longitudinal CF airway studies also showed strong concordance. For example, we achieved 96 % accuracy (24/25 samples correctly predicted) on strain-resolved metagenomics data (PRJEB32062)^27^. We achieved 74 % agreement (45/61 samples) in the bronchiectasis-CF comparison data, with most false negatives corresponding to culture-positive samples with extremely low *P. aeruginosa* read counts (PRJNA1055940)^28^. In larger contemporary cohorts, comprising more than 500 and 250 runs (PRJNA1081394 and PRJNA931830 respectively), our prediction accuracy exceeded 93 % and 99 %, matching reported culture trends before and after CFTR-modulator (ETI) therapy^29–31^.

Performance declined for oral swab datasets and early 454-based viromes, with uncertain provenance, when *P. aeruginosa* prevalence was genuinely low, or when metadata was unreliable. For example, oropharyngeal metagenomes (PRJNA1101448) yielded no predicted positives consistent with their report of only 2 % colonisation among adolescents^1^. In contrast, we correctly classified in 8 of 9 cases from matched sputum samples (PRJEB51171)^32^. Virome-dominated runs (PRJNA71831) and other legacy datasets could not be meaningfully evaluated^3^.

In summary, combining all datasets with usable metadata, 94% of runs (702 of 749 with confident provenance (Supplementary Tables 11 and 12) were correctly predicted for *P. aeruginosa* culturability. The few remaining discrepancies were attributable to short read lengths, limited coverage, or absence of contemporaneous culture data. The greatest deviations occurred in young paediatric cohorts with oral swab sampling, where *P. aeruginosa* prevalence is intrinsically low. Our results demonstrate that the South Australian–trained model generalises across sequencing platforms, geographies, and disease stages, accurately distinguishing *P. aeruginosa*-dominated airway microbiomes in CF metagenomes worldwide, and portend a future where these models can be used to predict the likely acquisition of *P. aeruginosa*.

## Discussion

High-dimensional biological datasets often exhibit complex interdependencies, where subtle correlations and redundancies among variables can obscure robust feature selection. We applied a random forest classifier to our short-read data in our initial analyses. However, across multiple runs, we observed substantial variability in feature importance rankings, with different variables emerging as the most predictive in multiple iterations. This instability suggests that the model is navigating a high-dimensional space with many near-equivalent solutions, where slight perturbations in data sampling or model initialisation resulted in shifts in feature selection. We tested metagenome assembled genomes, but the loss of discriminatory signals during the assembly process precluded the prediction of clinical outcomes. To address this challenge, we developed an entirely novel autoencoder-based approach for dimensionality reduction, distilling the high-dimensional dataset into a lower-dimensional latent space that captures essential biological variation while mitigating noise and redundancy. This structured representation ensured that we used a more consistent and interpretable feature set in our downstream classification, reducing variability between iterations of the same analysis and enhancing the robustness of our predictive framework.

The latent clusters derived from the autoencoder, which we call Clusters of Phylogeny and Functions (COPFs; pronounced *cough*), captured the dominant ecological and functional axes embedded within the CF airway microbiome. These clusters condensed thousands of taxonomic and metabolic features into a small number of interpretable “meta-features” that generalise across individuals, sequencing platforms, and time. The ordination of these COPFs revealed a continuous ecological gradient along our Medications Score. Samples with a low treatment burden and a low Medications Score were enriched in commensal genera, such as *Neisseria*, *Gemella*, and *Pasteurella*. In contrast, heavily treated samples were dominated by opportunistic genera, including *Achromobacter, Mycobacterium,* and *Pseudomonas*, and were enriched for canonical virulence functions such as lipopolysaccharide biosynthesis, siderophore production, and osmoprotectant metabolism. These patterns reflect what clinicians have known for years and define a disturbance–response continuum in which repeated therapy drives ecological restructuring rather than discrete shifts between metastable health and disease states.

A coherent signal emerged when we integrated community-level features with genome-resolved profiles. Across 127 samples, we identified 3,215 MAGs, 22 of which were annotated as *Pseudomonas* (at the genus level at least), and 14 of these were significantly enriched in *P. aeruginosa* culture–positive samples. This concordance indicates that genome-resolved fragments track the same biological signal as COPFs enriched for Pseudomonadaceae and for canonical *Pseudomonas*-associated functions (e.g., phenazine biosynthesis, *zot*-like elements). Notably, the majority of *Pseudomonas* MAGs are short, fragmented, and contain markers of the pan-genome; yet the abundance of reads mapping onto *Pseudomonas* MAGs is greater from samples from which *P. aeruginosa* was also cultured, suggesting that they represent genuine, strain-linked genomic partitions within the airway community rather than assembly artefacts. By contrast, analogous analyses found no reproducible associations between MAG abundance and either multidrug-resistant (*MDR*) phenotype or conversion to *P. aeruginosa* positivity within 12 months.

The COPFs derived from short-read taxonomy and functions capture broad axes of microbial adaptation, encompassing multiple co-existing *Pseudomonas* lineages and unbinned genomic fragments. In contrast, the *Pseudomonas* MAGs correspond to lineages within a highly polymorphic population. The weak correspondence between MAGs and the *Pseudomonas*-enriched COPFs highlights the collective nature of functional potential, whereby multiple taxa contribute overlapping genetic repertoires that define pathogenic phenotypes, and sequence assembly algorithms conflate the signatures of those taxa. Our latent dimensions constitute a higher-order abstraction that integrates shared functions across taxa, while the MAGs delineate the individual genomic contributors to that distributed network. Genome-resolved abundance alone thus incompletely describes the community-level roles of key pathogens in CF airways, emphasising the importance of functional co-variation as a lens through which to interpret pathogen ecology. Abundant microbes are critical, but less abundant microbes (those that cannot be assembled into complete genomes) create a conducive environment that enables pathogens to thrive.

By coupling COPFs with gradient-boosted random forests, we identified metabolic and respiratory modules that stratify multidrug-resistant (MDR) and non-MDR *P. aeruginosa* isolates. MDR-associated clusters were depleted in NADH:quinone oxidoreductase and menaquinone biosynthesis genes but enriched for redox-buffering and anabolic functions, including Calvin–Benson cycle enzymes and pyrimidine biosynthesis. These patterns mirror experimental findings that disruption of respiratory-chain components promotes biofilm formation, virulence, and antibiotic tolerance. It is perhaps surprising that the changes observed *in vitro* are also observed *in homine*. Together, they suggest that metabolic rewiring rather than canonical resistance genes underpins much of the multidrug-resistance phenotype, consistent with recent models of physiological plasticity driving antibiotic resilience in chronic *P. aeruginosa* infections^33,34^.

Beyond established infection, our latent-feature framework predicts impending *P. aeruginosa* colonisation up to a year in advance. The capacity to spae pathogen emergence transforms metagenomics from a retrospective diagnostic into a prospective clinical tool, offering physicians a critical window to prevent chronic infection before it becomes irreversible. The COPFs enriched in individuals who later converted to culture positivity collectively signal a shift toward a pathogen-permissive airway environment. Conversion risk was not defined by the emergence of a single precursor species, but by a distributed microbial remodelling across the community. We propose that airway microbial ecosystems transition through functionally altered intermediate states that predispose to pathogen takeover, and using metagenomics, we might be able to provide an early-warning signal of clinical decline (Fig. 6).

**Fig. 6.**
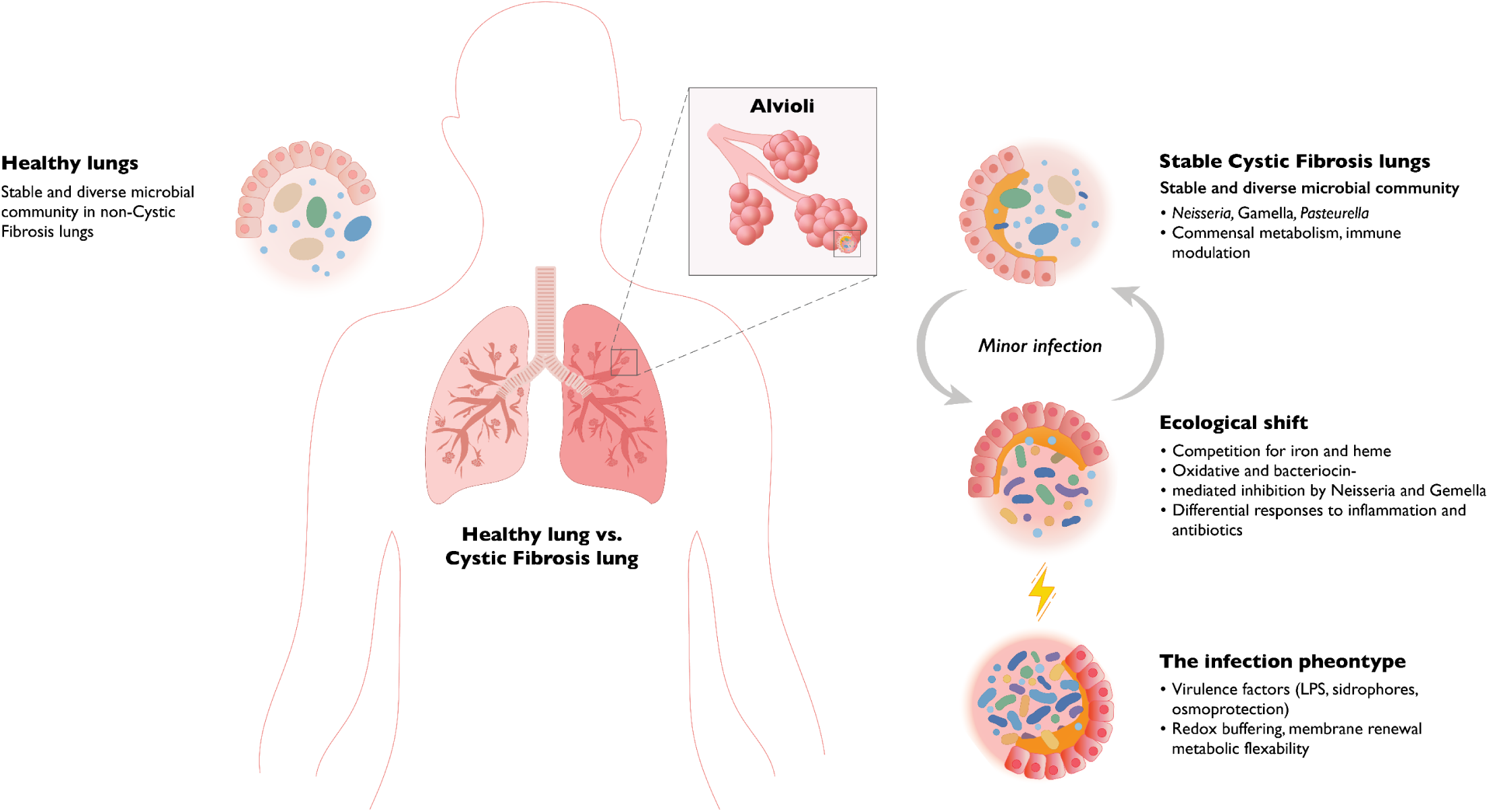
Conceptual model for infection progression in pwCF. Repeated infections destabilize the commensal microbial flora, cause damage through inflammation, and the administration of antibiotics results in the development of the infective phenotype.

Metagenomics is not a panacea for detecting all pathogens. We demonstrated that culturing remains essential for some taxa, notably *S. maltophilia*, that continue to plague the CF airway. However, by incorporating eukaryotic markers, our framework extended predictive metagenomics to encompass fungal ecology. Although antifungal therapy and spatial heterogeneity limit the sensitivity of culture-based diagnostics, metagenomic sequencing consistently detected persistent fungal DNA even during culture-negative intervals. Whether this reflects under-detection by culture or over-permissive detection by metagenomics remains unresolved, underscoring the need for integrated molecular and culture-based benchmarks in future clinical mycobiome studies. We note that ABPA lacks a universally accepted gold-standard clinical definition. Contemporary ISHAM criteria integrate multiple partially sensitive markers, meaning that diagnostic uncertainty is intrinsic to ABPA. This imprecision constrains the evaluation of metagenomic models, in which ambiguity in the clinical label may overshadow the true biological signal.

To assess generalisability, we applied our South Australian–trained classifier to >1,500 publicly available CF metagenomes from 22 BioProjects worldwide. Despite spanning multiple sequencing technologies, sample types, samples pre- and post-modulator therapy, and geographic origins, the model achieved 94 % accuracy in predicting *P. aeruginosa* culture results. Errors predominantly arose in datasets with low read depth, single-end sequencing, or uncertain sample provenance, particularly oropharyngeal swabs from paediatric cohorts with intrinsically low *P. aeruginosa* prevalence. The consistency of predictions across independent datasets indicates that the latent ecological and functional axes captured by COPFs are globally conserved within CF microbiomes.

Although our cohort largely reflects the pre-modulator era, we recognise that the widespread adoption of ETI is reshaping the clinical and microbial landscape of cystic fibrosis. We therefore acknowledge this as a limitation, but emphasise that our dataset provides an essential baseline against which post-modulator airway microbiomes can be compared. Together, our findings redefine the CF airway microbiome as a dynamic ecological continuum rather than a binary landscape of infection, underscoring the importance of incorporating ecological interactions and resulting gene function capacity in the microbiome for informed predictions. The ability to predict pathogen presence, resistance, and impending colonisation from metagenomic community structure establishes a foundation for culture-independent diagnostics. These results also provide an ecological interpretation of increased rates of antibiotic resistance. COPF-derived latent dimensions summarise complex ecological and functional relationships into interpretable markers that can be integrated into clinical workflows, complementing traditional culture and molecular assays. Beyond CF, this approach demonstrates how ecological theory and deep learning can transform metagenomics from descriptive surveys into predictive, mechanistic models of disease progression.

## Methods

### Sample Collection

This study was approved by the Human Research Ethics Committee and Women’s and Children’s Health Network (HREC/16/WCHN/30). In total, 127 sputum samples were collected during routine outpatient and inpatient clinical care of 64 pwCF at the Women’s and Children’s Hospital and Royal Adelaide Hospital between 2017 and 2019. All were standard, clinically indicated samples (either spontaneous expectoration or induced) that were divided: approximately 0.5-1 mL of sputum was reserved for metagenomic analysis and stored at −80°C until analysis, with the remainder sent to SA Pathology for routine testing. The treating clinician determined each participant’s clinical status at the time of sampling. Clinical metadata, including FEV_1_/best FEV_1_ and prescribed medication, were also collected.

### Sample preparation and DNA extraction

After thawing at room temperature, 0.5-1 mL sputum aliquots reserved for metagenomic sequencing were adjusted to a final volume of 4 mL using sterile, autoclaved, 1X Phosphate Buffered Saline (PBS). The samples were homogenised using a 1 mL syringe until no visible sputum clumps remained, then divided into two equal 2 mL aliquots: one for microbial metagenomics and one for viral metagenomics. This manuscript only used the 2 mL aliquot reserved for microbial metagenomics.

Microbial DNA was extracted immediately from the sputum using methods previously described^3^. Briefly, the 2 mL of homogenised sputum reserved for microbial metagenomics was further diluted 1:5 with sterile 1x PBS, followed by the addition of β-mercaptoethanol (2% v/v) to reduce sputum viscosity. The mixture was incubated at room temperature on a rocking platform for 2 hours, then centrifuged at 3,056 x g for 15 minutes at 10°C. The resulting pellet was resuspended in 10 mL of nuclease-free water and incubated for 15 minutes at room temperature to lyse the host cells selectively, then centrifuged again at 3,056 x g for 15 minutes at 10°C. This process of host cell lysis was repeated twice. After the second wash, the pellet was resuspended in 5 mL of 1X DNase buffer (50 mM NaAc, 10 mM MgCl2, 2 mM CaCl2; pH 6.5, and host DNA was subsequently depleted by treatment with 75 µL DNase I (Sigma-Aldrich, cat# D5025-15KU) and incubation at 37°C for 2 hours with repeated mixing. The DNaseI-treated sample was then centrifuged again at 3,056 x g for 15 minutes at 10°C, and the bacterial pellet was washed twice in SE buffer (75 mM NaCl, 25 mM EDTA; pH 7.5) to inactivate DNaseI activity. The final pellet was resuspended in 1 mL of SE buffer, transferred to a microfuge tube, and centrifuged at 16,100 x g at room temperature for 15 minutes. The supernatant was discarded and the DNA extracted from the pellet using the NucleoSpin® Tissue Kit (MACHEREY-NAGEL, cat#: 740952.50) following the ‘hard-to-lyse bacterial (Gram-positive)’ protocol. DNA was eluted from the spin column in the final step of the protocol using two separate elutions. For each elution, 35 µL of elution buffer (5 mM Tris/HCl, pH 8.5; provided with the kit) was added directly to the spin column membrane, incubated at room temperature for 3 minutes, and then centrifuged at 11,000 x g for 1 minute. Eluates were collected into separate 1.5 mL tubes (i.e. one tube containing 35 µL from the first elution and another tube containing 35 µL from the second elution). DNA concentrations from the first elution were measured using the Qubit 1x dsDNA High Sensitivity Assay Kit (Invitrogen), with yields ranging from undetected concentrations (< 0.0005 ng µl^−1^) on the Qubit to 18 ng µl^−1^.

### Library preparation and DNA sequencing

Extracted DNA was sequenced using short-read MGI sequencing at the South Australian Genomics Centre (Adelaide, South Australia) or long-read Oxford nanopore sequencing using the MinION and PromethION sequencing platforms in-house (Flinders University, FAME lab).

For short-read sequencing, mechanical shearing via sonication was performed on 50 µL of eluate, using the Qsonica Q800R (Qsonica, USA). Sonication targeted a 650 bp length (2×300bp); amplitude 20% for 12 minutes total time (6 minutes active sonication), alternating 15 sec on and off. Library preparation was performed using the xGen™ 2S DNA Library Prep kit (Integrated DNA Technologies, USA) following the manufacturer’s protocol for ‘less than 10ng gDNA’ with 12 cycles in the indexing PCR. Libraries were sent to the South Australian Genomics Centre, converted from linear DNA into the required MGI nanoball structures using the MGIEasy kit (MGI, China), and sequenced on the MGI DNBSEQ-G4000, generating 300 bp reads. All 127 samples were prepared and sequenced using short-read MGI technology.

For MinION and PromethION long-read sequencing, the Rapid PCR barcoding kit (SQK-RPB004 for MinION; SQK-RPB114.24 for PromethION; Oxford Nanopore) was used according to the manufacturer’s instructions with the following modifications. For the PCR amplification step, the number of PCR cycles was increased from 14 to 25. PCR products were concentrated using SPRIselect beads (Beckman Coulter) for MinION samples and AMPure XP beads for PromethION samples. To maximise DNA yield during the final elution step, magnetic bead-bound DNA was incubated at 45°C for 15 minutes in sterile, nuclease-free water for SPRIselect bead preparations or in Elution Buffer (EB) for AMPure XP bead preparations. Concentrated PCR product was quantified using the Qubit 1X dsDNA High Sensitivity Assay Kit (Invitrogen) before sample pooling and loading onto the MinION or PromethION. Following 25 PCR cycles with the SQK-RPB004 kit, DNA concentrations for the 61 samples sequenced on the MinION ranged from 2 to 200 ng µl^−1^. For the 10 samples prepared using the SQK-RPB114.24 kit, DNA concentrations ranged from 0.8 to 41 ng µl^−1^ after 25 PCR cycles. Although DNA concentrations were lower in samples prepared for PromethION sequencing, the PromethION platform provides a greater sequencing depth than the MinION and was still able to capture the microbial community composition (Supplementary Fig. 1). Samples were sequenced using the R9.4.1 flow cell chemistry on the MinION and the R10.4.1 flow cell chemistry on the PromethION (Oxford Nanopore). A total of 61 samples from 43 pwCF were sequenced using the MinION, while 10 samples from 9 pwCF were sequenced using the PromethION. The 10 samples selected for sequencing on the PromethION were chosen based on having the highest DNA concentrations from the first DNA elution step. Where less than 3 µL were available from the first elution, the second elution for that sample was used to make up the required volume.

### Long- and Short-Read Metagenome Annotations

Each metagenomics sequencing run was processed separately using a very similar pipeline. All computations were performed on Flinders HPC Deepthought^35^ or Pawsey’s HPC, Setonix^36^. The primary difference is that short-read sequencing was paired-end and the paired-end reads were processed together, while long-read sequencing was unpaired and processed singly. In the following commands, the variable $READ is used to indicate a specific run name for long read sequences and $R1 and $R2 are used to indicate the R1/R2 paired-end runs.

The long-read data were cleaned with *fastp* (version 1.0.1)^37^ using the parameters:

~~~
fastp -n 2 -l 50 -i fastq/$READ -o fastq_fastp/$READ --adapter_fasta
∼/atavide_lite/adapters/ONT_RPB114.24.fna
~~~

The *fastp* filtering removes sequences with two or more “N” bases, removes the ONT adapters, and requires a minimum read length of 50 bp, effectively removing low-quality Nanopore reads.

The short-read data were cleaned with *fastp* (version 1.0.1)^37^ using the parameters:

~~~
fastp -n 1 -l 100 -i fastq/$R1 -I fastq/$R2 -o fastq_fastp/$R1 -O fastq_fastp/$R2 -j
fastp_output/$R1.json -h fastp_output/$R1.html --adapter_fasta
atavide_lite/adapters/IlluminaAdapters.fa --thread 16
~~~

For the short reads, we remove sequences with a single “N” base, sequences shorter than 100 bp, and trim the Illumina adapters.

Next, the sequences were compared to the human genome reference sequence GRCh38 using minimap2 (version 2.30-r1287)^38^ and samtools version 1.21^39^ with the following command for long read sequences:

~~~
minimap2 -t 64 --secondary=no --split-prefix=tmp$$ -a -x map-ont GRCh38.fna
fastq_fastp/$READ | samtools view -bh | samtools sort -o $READ.bam –
~~~

And the similar command for short read sequences:

~~~
minimap2 -t 16 --split-prefix=tmp$$ -a -xsr $HOSTFILE $QC/$R1 $QC/$R2 | samtools view
-bh | samtools sort -o host_bamfiles/$R1 -
~~~

These commands use the default *minimap2* parameters for mapping reads against a reference genome and have been optimised by the tools’ authors^38^.

The sequences were identified from the BAM file created by the previous command and filtered into two separate files using samtools (both *samtools* and *htslib* version 1.22.1)^39^. Sequences that do *not* match the flag 3588, and thus represent mapped reads that pass the platform/vendor quality check, are not PCR or optical duplicates, and are primary alignments, were filtered as mapping to the human genome. For short reads, additional samtools flags were used to identify R1 reads (matching flag 65) and R2 reads (matching flag 129). Reads that do *not* match the flag 3584, and thus represent unmapped reads which pass the platform/vendor quality check, are not PCR or optical duplicates, and are primary alignments, are filtered as unmapped reads, and again, short reads were separated using flags that match 77 for R1 reads and 141 for R2 reads.

Reads that did not map to the human genome were mapped against the UniRef50^40^ database using MMseqs2 (version 18.8cc5c) with the following command:

~~~
mmseqs easy-taxonomy fasta/$R1 $DBSOURCE/$DB $OUTPUT/$TMPOUTPUT $TPD --threads 32
~~~

The authors optimised MMseqs2’s easy-taxonomy parameters for a balance of sensitivity and speed, including a 7-mer double-match prefilter with compositional bias correction, low-complexity masking with TANTAN, and default similarity thresholds (--k-score ∼95) that generate sufficient similar *k*-mers for accurate detection while maintaining computational efficiency. Candidate hits are further filtered by fast ungapped alignment and refined with vectorised Smith–Waterman alignments, after which taxonomic labels are assigned using a lowest common ancestor (LCA) approach described in more detail in their paper and on their website.

The distribution of MMseqs2 alignment E-values spanned a wide range, from highly significant matches (minimum E-value of 0) to weak or non-significant hits (maximum 0.9). The bulk of matches were strongly significant, as reflected by a median of 6.2 × 10⁻¹⁸, with the average skewed upward (1.4 × 10^−3^) by a minority of higher E-values. These summary statistics indicate that most alignments were highly significant, while a small fraction of weak matches contributed disproportionately to the mean.

The taxonomies were reformatted with TaxonKit (version 0.20.0)^41^ and merged into a single table.

The functional annotations were enriched by directly mapping UniProt IDs from the MMseqs2 output to proteins associated with BV-BRC subsystems (a 1:1 mapping that did not require thresholds/cutoffs)^42^. The number of reads MMseqs2 reported as mapping to each protein was counted, and the total was divided by the number of mapped reads to normalise the read counts.

For the assembly-based approaches, the metagenomes were assembled using *MEGAHIT* (version 1.2.9)^43^, which by default employs a succinct de Bruijn graph approach with multiple *k*-mer sizes (21 to 141), automatic memory optimisation, and conservative heuristics for contig pruning to balance assembly sensitivity, contiguity, and computational efficiency. Metagenome-assembled genomes (MAGs) were reconstructed using VAMB (version 5.0.4)^44^. This variational autoencoder-based approach jointly encodes *k*-mer composition and contig differential coverage profiles into a low-dimensional latent space. By learning these compressed representations, VAMB effectively separates contigs originating from different microbial genomes and clusters them using an iterative medoid clustering algorithm. This deep learning approach has been shown to improve bin purity and completeness compared to conventional binning methods such as MetaBAT2^45^ or CONCOCT^46^, particularly in complex metagenomic datasets.

### Assembly Bins and Metagenome Assembled Genomes (MAGs)

MAGs were constructed using cross-assembly and VAMB^44^. Briefly, all long- and short-read data were combined and assembled using MEGAHIT^43^. Assembly took approximately 16 days to complete, using 272.45 GB of RAM and 200 compute cores. All reads were mapped back to all contigs using minimap2, and the contigs were then binned using VAMB. Sequence coverage was calculated using Koverage^47^. For each bin, the median sequence read coverage was calculated from all contigs mapping to that bin. Metagenome-assembled genome bins were annotated using the BV-BRC command-line annotation system (the “p3-scripts”). Taxonomic assignment was performed using the BV-BRC Mash “compute_genome_distance_for_fasta2” service. For each genome, the top mash hit was used to identify the genus.

### Bacterial Nomenclature

Clinical culture reports typically describe the actual bacterial genus and species (e.g., *P. aeruginosa, S. maltophilia*). In contrast, the taxonomic assignments described above usually annotate at the family (e.g., Pseudomonadaceae, Lysobacteraceae) or genus (e.g., *Pseudomonas, Stenotrophomonas*) levels. Throughout, we have used the most appropriate nomenclature for the available classification level.

### Medications Score

To derive an integrated quantitative measure of overall clinical health, we combined metadata variables related to medication and treatment burden. These included whether each pwCF was prescribed antibiotics, the total number of antibiotics, total number of medications, total number of antifungals, and the total number of steroids or monoclonal antibodies at the time of sampling. We applied principal component analysis (PCA) to these five variables using the scikit-learn (version 1.6) implementation in Python (version 3.12)^48^. The first principal component captured the dominant axis of variation across all treatments and was used as a composite “Medications Score”. This Score was linearly scaled from 0 to 100, with higher values corresponding to greater medication load and reduced clinical stability. The Medications Score was subsequently used in downstream analyses to evaluate associations between microbiome composition, predicted virulence potential, and overall patient health.

Because pwCF experience lifelong decline in lung function, raw or normalised FEV_1_ measures are less appropriate for identifying lung-function distress. Therefore, we use the ratio of the current FEV_1_ to the best FEV_1_ recorded in the last 12 months. We also categorise it as an FEV_1_ ratio score: 1 if the ratio is 0.95 or higher, 2 if the ratio is between 0.9 and 0.95, and 3 (the worst) if the ratio is less than 0.9.

### Allergic Bronchopulmonary Aspergillosis

We used immunological markers routinely measured in pwCF to identify *A. fumigatus* sensitisation or allergic bronchopulmonary aspergillosis (ABPA). Across our cohort, total IgE ranged from 0 - 1,750 IU mL⁻¹, *A. fumigatus*-specific IgE from 0–61.5 kUA L⁻¹, specific IgG from 4–200 mgA L⁻¹, and precipitin titres from 0 (negative) to 2 (strongly positive). Following the ISHAM consensus criteria^24^, we defined *Aspergillus* sensitisation as *A. fumigatus*–specific IgE ≥ 0.35 kUA L⁻¹ and total IgE < 500 IU mL⁻¹. We defined ABPA as *Aspergillus* sensitisation (specific IgE ≥ 0.35), and total IgE ≥ 500 IU mL⁻¹ and either specific IgG ≥ 27 mg A L⁻¹ or positive precipitins (our metadata includes neither radiology nor eosinophilia, which are also used as supportive criteria).

### Clusters of Phylogeny and Functions (COPFs)

We analysed 127 samples from 64 pwCF to predict 166 clinical outcomes. However, our taxonomic profiles ranged from kingdom (1 group) to genus (3,581 groups), while functional annotations were characterised at multiple subsystem levels from class (11 features) to functions (6,230 features). We trained a deep, fully connected, undercomplete autoencoder (with a 50-dimensional bottleneck) to cluster the data, preventing overfitting and mitigating data imbalance, which is particularly critical given our sample size. The autoencoder was built in Python using TensorFlow/Keras (version 2.13.1)^49^.

Our autoencoder started with the same input dimensions as the number of variables. Unless otherwise described, for the analysis presented here, we trained an autoencoder using family taxonomic level (743 features) and subsystem-level functional annotations (769 features) with 1,512 input dimensions. We used two layers of densely connected neural networks, first with 512 units and the second with 128 units, separated by a 20% dropout layer. This autoencoder architecture was chosen as a compact, non-linear model that balances flexibility with overfitting risk for our dataset of 127 samples and ∼1,500 features. Layer widths were selected from a small set of candidate sizes and retained because they yielded low validation loss under early stopping and stable latent clusters for downstream analyses. Finally, our encoder ended with 50 latent dimensions. Each layer used the rectified linear unit (ReLU) activation function. We trained the autoencoder using the Mean Squared Error (MSE) loss function and the Adam optimiser until the value loss stopped improving. Early stopping was triggered after 15 epochs without improvement in validation MSE, and the best weights were restored. For the model with 1,512 input dimensions, training took 54 epochs. To identify groups of covarying taxonomic and functional features, we computed Pearson correlations between each input variable and the 50 latent dimensions of the trained autoencoder. The resulting correlation matrix (1,512 × 50) captures the degree to which each microbial feature covaries with each latent dimension, effectively linking compositional and functional variation to abstract representations learned by the neural network. We then applied hierarchical agglomerative clustering using Ward’s linkage and Euclidean distance to the feature–latent correlation profiles, resulting in a dendrogram that grouped features by their shared correlation structure. The dendrogram was cut at *k* = 150 to generate compact sets of features with similar correlation signatures. We selected *k* = 150 as a pragmatic balance between resolution and stability, given the dimensions of our dataset. This selection corresponds to compressing the feature space by roughly an order of magnitude, yielding clusters large enough to capture correlated biological signals while avoiding over-fragmentation of the data. Pilot runs using coarser and finer cuts (e.g., ±25–50% of this value) produced highly similar downstream results, indicating that the precise choice of *k* does not strongly affect cluster-level patterns.

Each resulting cluster, which we named a “Cluster of Phylogeny and Function” (COPF) and represents a coordinated axis of taxonomic and functional variation, was subsequently used as a meta-feature in downstream analyses and predictive models, including our gradient-boosted random forests.

### Gradient-Boosted Random Forest

All models were implemented in Python using scikit-learn (version 1.6)^8,48^. Metadata variables were either numeric (e.g., age) or categorical (e.g., inpatient vs. outpatient). Categorical predictors and outcomes were encoded with the OrdinalEncoder. For each outcome, the dataset was randomly partitioned into an 80% training set and a 20% held-out test set. Gradient-boosted forests were restricted to a conservative parameter regime tailored to the structure of our dataset. We trained up to 1,000 boosting iterations to limit the number of candidate splits and reduce variance; trees were capped at a depth of 5 with a minimum of 5 samples per leaf, and the learning rate was set to 0.005. These settings were chosen to balance model flexibility with overfitting risk, and reflect common practice for high-dimensional, small-sample biological datasets. Exploratory runs with moderately larger or smaller learning rates, depths, and numbers of estimators produced similar feature-importance profiles, so we retained the more constrained configuration to ensure stable feature ranking. Within the training data, 20% was held out for internal validation, and early stopping was applied if validation loss failed to improve for 20 consecutive boosting iterations (Supplementary Table 13). When early stopping was triggered, the effective number of estimators was smaller than 1,000. Final performance was assessed on the untouched 20% test set: mean squared error was used for regression tasks, whereas for binary outcomes predicted probabilities were used to compute receiver–operating characteristic (ROC) curves and the area under the ROC curve (AUC). Variable importance was extracted from the fitted models as Gini impurity–based feature importances and visualised using seaborn (version 0.13) and matplotlib (version 3.8). All code for creating the ensemble gradient-boosted random forest and for plotting variable importance is available in our GitHub repository^8^.

### Worldwide CF Sample Analysis

Samples were identified by searching the SRA using NCBI and Google’s BigQuery. First, the NCBI SRA was searched for any BioProject with the phrase “Cystic Fibrosis” (available at https://www.ncbi.nlm.nih.gov/bioproject/?term=cystic+fibrosis). In February 2025, this resulted in 1,187 BioProject IDs. Next, the SRA metadata was queried using Google’s BigQuery. First, we built a temporary table that contains all entries where either the assay type is “AMPLICON” or the library selection is “PCR”. Next, we selected entries in the SRA metadata where the library source is “METAGENOMIC” or “METATRANSCRIPTOMIC”, or the organism contains “microbiom” or “metagenom” (to find microbiome/microbiomics or metagenome/metagenomics and other variants). We cross-referenced the BioProjects from this list with those from the SRA search for “Cystic Fibrosis” and excluded any BioProjects that were entirely based on amplicon sequencing. Finally, we filtered the dataset for the “organism” field to contain one or more of the following entries: *human lung metagenome; metagenome; human sputum metagenome; Homo sapiens; human nasopharyngeal metagenome; lung metagenome; human metagenome; mixed culture metagenome; clinical metagenome; or respiratory tract metagenome*. The last filtering step largely (but not entirely) eliminates the gut microbiome samples. Individual fastq files were downloaded using Kingfisher, and then each run was processed using the atavide-lite pipeline, as described above. Data was imported into Jupyter notebooks available on GitHub for processing and analysis. Samples with uncertain provenance are those for which the publicly available SRA metadata could not be confidently matched to the published clinical or culture records, or where the sequencing type (e.g., single-end, 454, or virome-only) or read quality prevented verification.

## Data Availability

All data were analysed using Python (versions 3.11 and higher), and a complete repository of the processed data is available at https://github.com/linsalrob/CF_Data_Analysis. The sequence data is available from the EGA under Data Access Policy EGAP50000000796, Study Accession Number EGAS50000001488, and run accessions EGAR50000337887-EGAR50000338084.

## Ethics Approval

The samples were collected under ethics approval from the Women’s and Children’s Hospital Network Human Research Ethics Committee approval number HREC/16/WCHN/30. The study was conducted in accordance with the principles of the Declaration of Helsinki (1964) and its subsequent amendments. All participants or their guardians provided informed consent for the collection of samples and the use of data in this study. The pwCF identifier is a study-specific pseudonym that is not derived from, and has no linkage to, any hospital or medical record identifier; re-identification is not possible within the research dataset and can only be performed by the treating clinician overseeing the study.

## Author Contributions

TRG conceptualisation, Data curation, Formal analysis, Investigation, Methodology, Project administration, Resources, Supervision, Writing – original draft, Writing – review & editing. JAPCJ data curation, Formal analysis, Investigation, Methodology, Resources. JM investigation, Supervision. CYO, IESE, JRI, MPD, PGS writing – review & editing. AT, AnT, MJR, SRG formal analysis. MSW methodology, Supervision, Investigation. BM, JW methodology. EH, HPAJ, SDP investigation. KLW writing – review & editing, Formal analysis. GB data curation, Formal analysis, Visualisation. NWF, RG visualisation. VM writing – review & editing, Software, Formal analysis. FJR formal analysis, Data curation. BP data curation, Formal analysis, Methodology, Software. AH, ALKH, BD, CK, CL, CMH, ENK, PD, QY, RDH, RJH investigation, Formal analysis. SKG project administration, Resources, Supervision. LKI data curation, Software. JGM writing – original draft, Writing – review & editing. CDW data curation, Investigation. EAD methodology, Funding acquisition, Conceptualisation, Formal analysis. RAE conceptualisation, Data curation, Funding acquisition, Project administration, Writing – review & editing.

## Funding Acknowledgements

MJR acknowledges funding from NHMRC Ideas Grant 2036844. BD acknowledges funding from the Cooperative Research Centre for Solving Antimicrobial Resistance in Agribusiness, Food, and Environments (CRC SAAFE). PGS acknowledges funding from the National Health and Medical Research Council of Australia (Idea Grants 2020173 and 2036550). RAE acknowledges funding from the NIH NIDDK RC2DK116713 and awards from the Australian Research Council DP220102915, DP250103825, and FL250100019. This research was supported by the Australian Government’s National Collaborative Research Infrastructure Strategy (NCRIS), with access to computational resources provided by the Pawsey Supercomputer Centre through the National Computational Merit Allocation Scheme, and DNA sequencing supported by Bioplatforms Australia, provided by the South Australian Genomics Centre.

---

**Supplementary Fig. 1.**
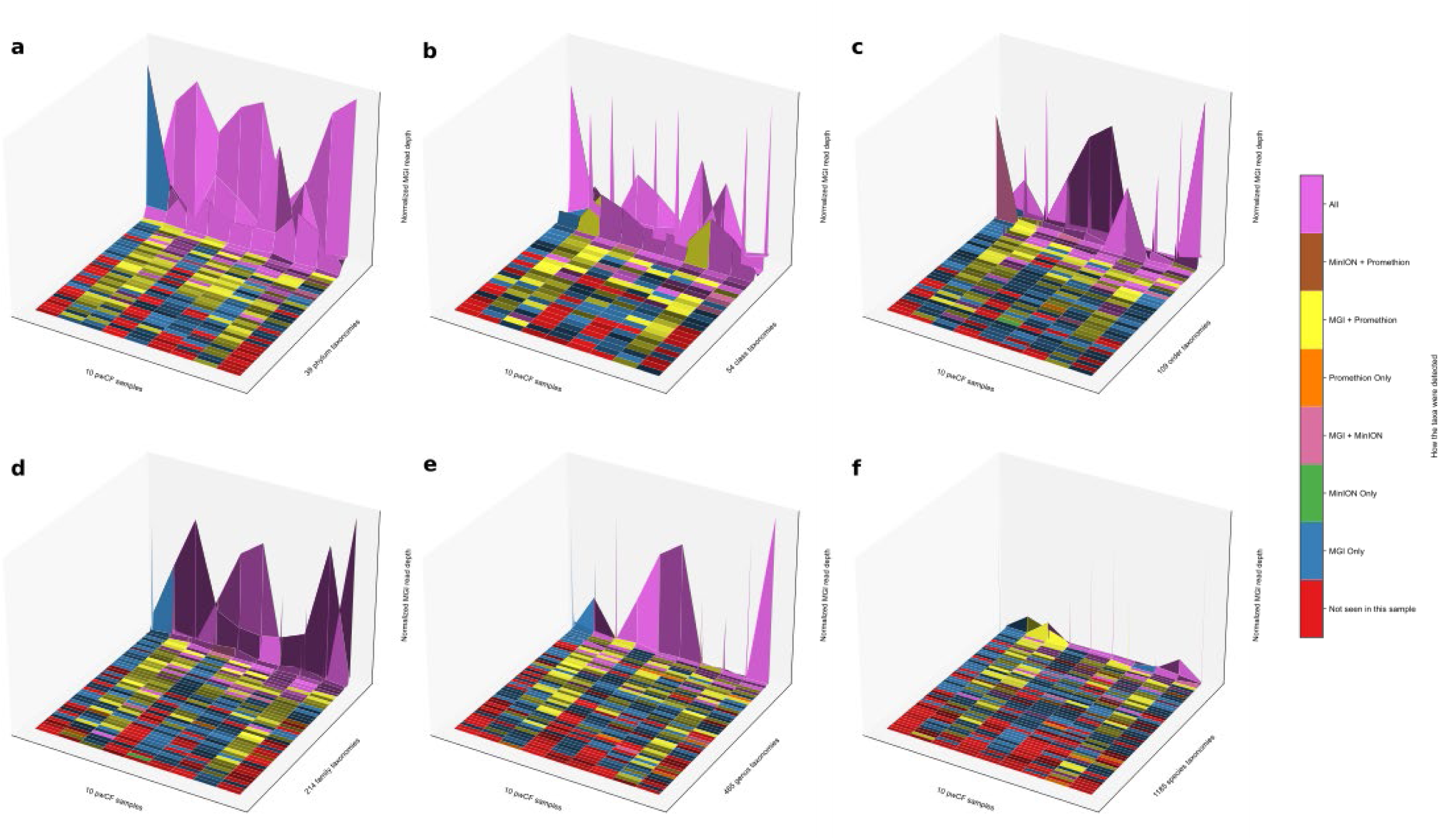
Comparison of species identified using MGI short read, Oxford Nanopore Minion, and Oxford Nanopore PromethION sequencing. The *n =* 10 samples that were sequenced using all three technologies are shown on the x-axis, the different taxa (unlabelled) are shown on the y-axis, and the normalised MGI read abundance is shown on the z-axis. Only those taxa with at least 10 reads per million mapped reads are shown. Colours indicate whether the taxa was identified with any of the sequencing technologies. Each panel reflects a different taxonomic rank: a. 39 phyla, RPKM ranges up to 811,485; b. 54 classes, RPKM ranges up to 781,208; c. 109 orders, RPKM ranges up to 694,463; d. 214 families, RPKM ranges up to 636,830; e. 465 genera, RPKM ranges up to 597,643; f. 1,185 species, RPKM ranges up to 388,863.

**Supplementary Fig. 2.**
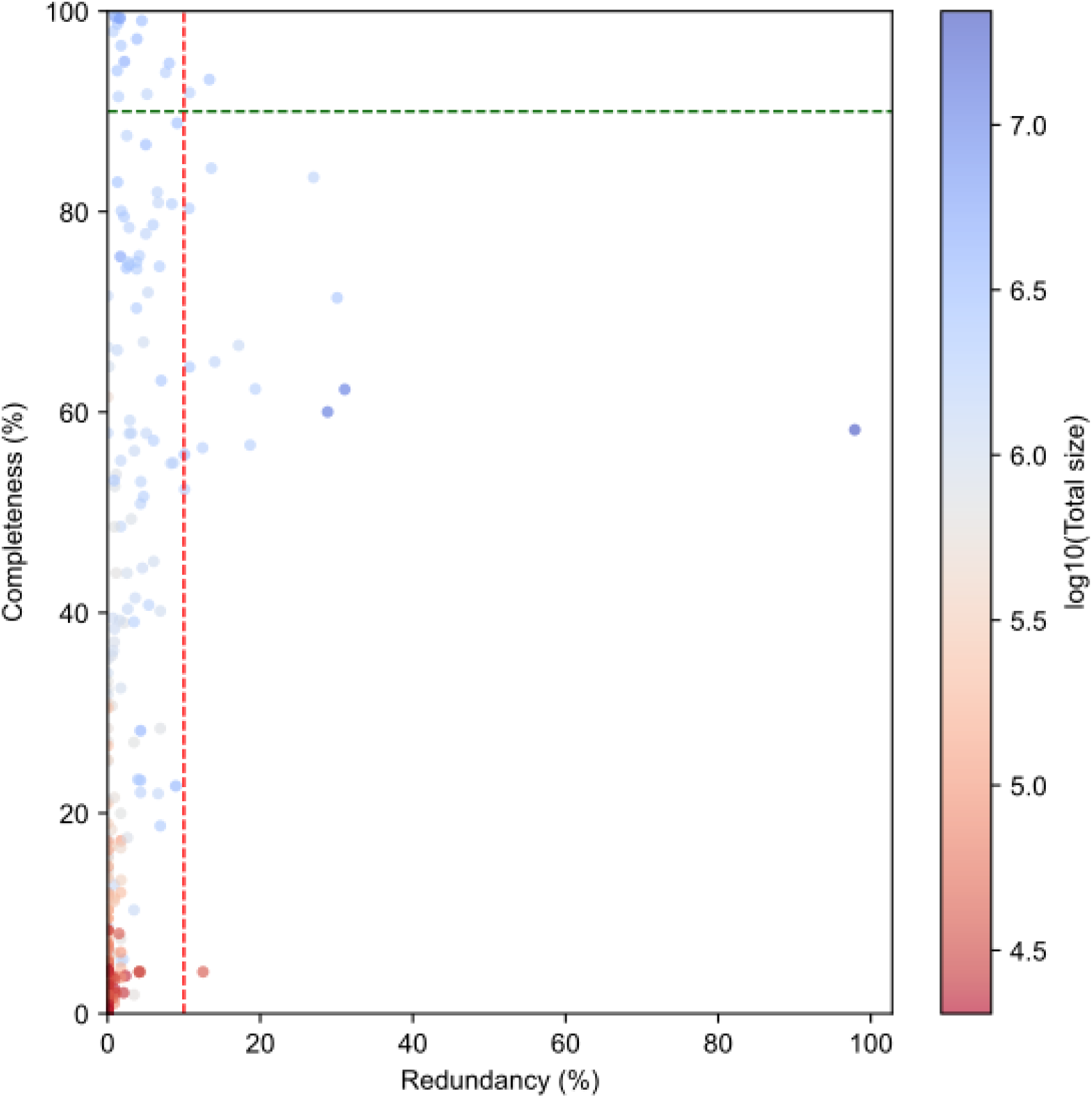
Relationship between completeness, redundancy, and total assembled genome size across all metagenome-assembled genomes (MAGs). Each point represents a MAG, positioned by its redundancy (x-axis) and completeness (y-axis), and coloured by the log_10_ of its total assembly size. Dashed lines mark thresholds commonly used to define high-quality metagenome-assembled genomes (≥90% completeness and ≤10% redundancy).

**Supplementary Fig. 3.**
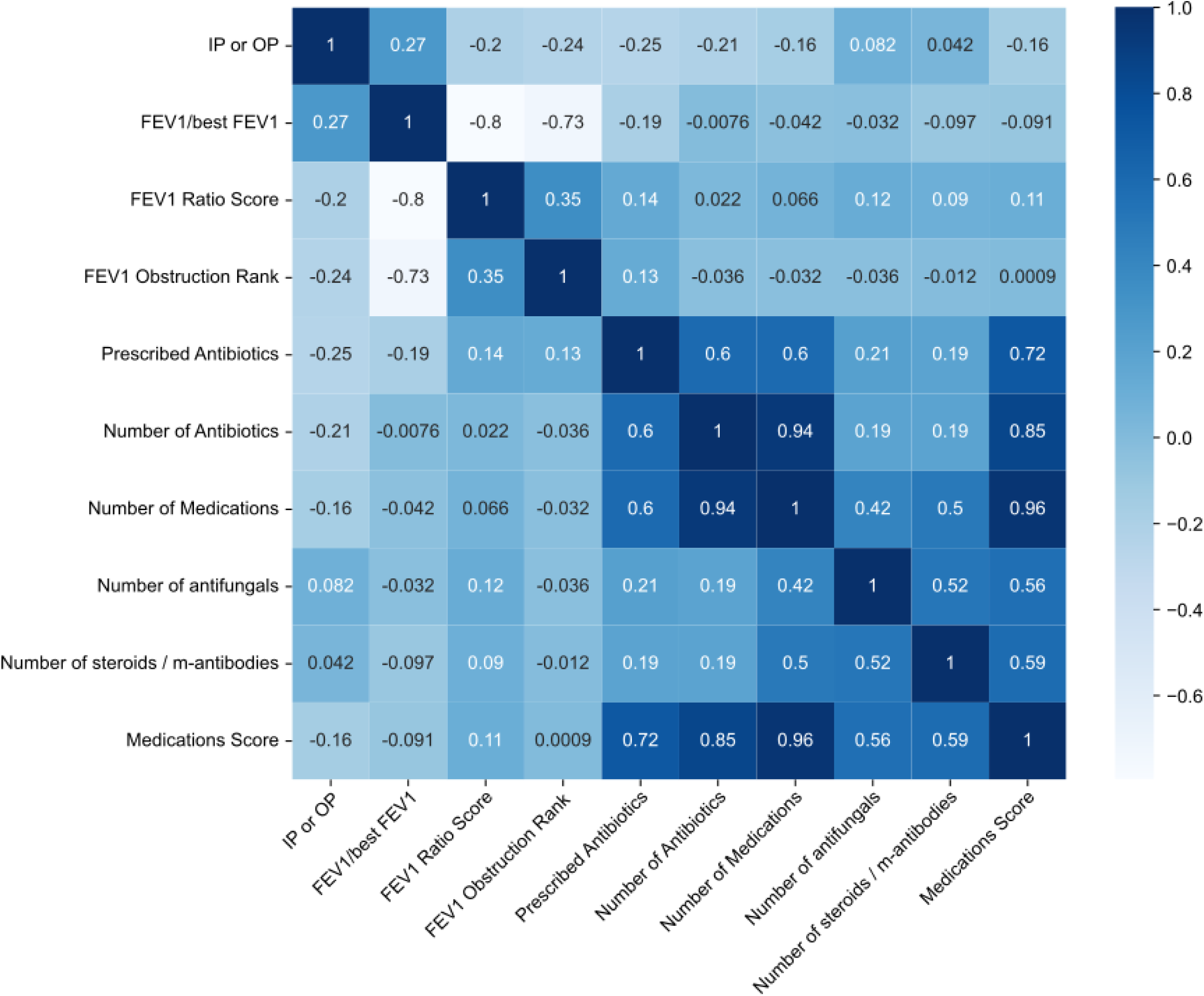
Pearson correlation of Medications Score and independent clinical measures. Pearson correlation matrix of the Medication Score and independent clinical measures. Each box is shaded by the strength and direction of the linear correlation (Pearson r), with the corresponding r-value shown inset (*n* = 125).

**Supplementary Fig. 4.**
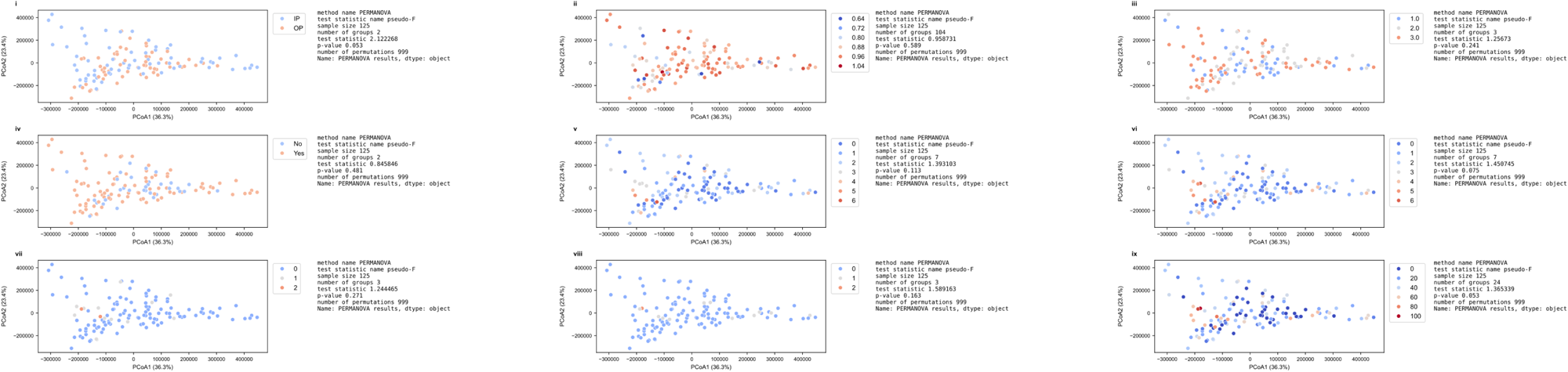
PCoA of the CPAF coloured by health measure. Each plot is the same scatterplot; however, we have coloured each point by the respective health score. We used PERMANOVA to calculate the clustering’s statistical significance (pseudo-*F*) shown to the right of the figure. i) in-patient vs outpatient (p=0.07); ii) FEV_1_/best FEV_1_ (p=0.96); iii) FEV_1_ ratio score (p=0.22); iv) Whether they were prescribed antibiotics (p=0.53); v) How many antibiotics they were prescribed (p=0.12); vi) How many medications they were prescribed (p=0.08); vii) How many antifungals they were prescribed (p=0.26); vii) How many steroids and monoclonal antibodies they were prescribed (p=0.17); ix) Medications Score (p=0.03**) (*n*=125 for each panel)

**Supplementary Fig. 5.**
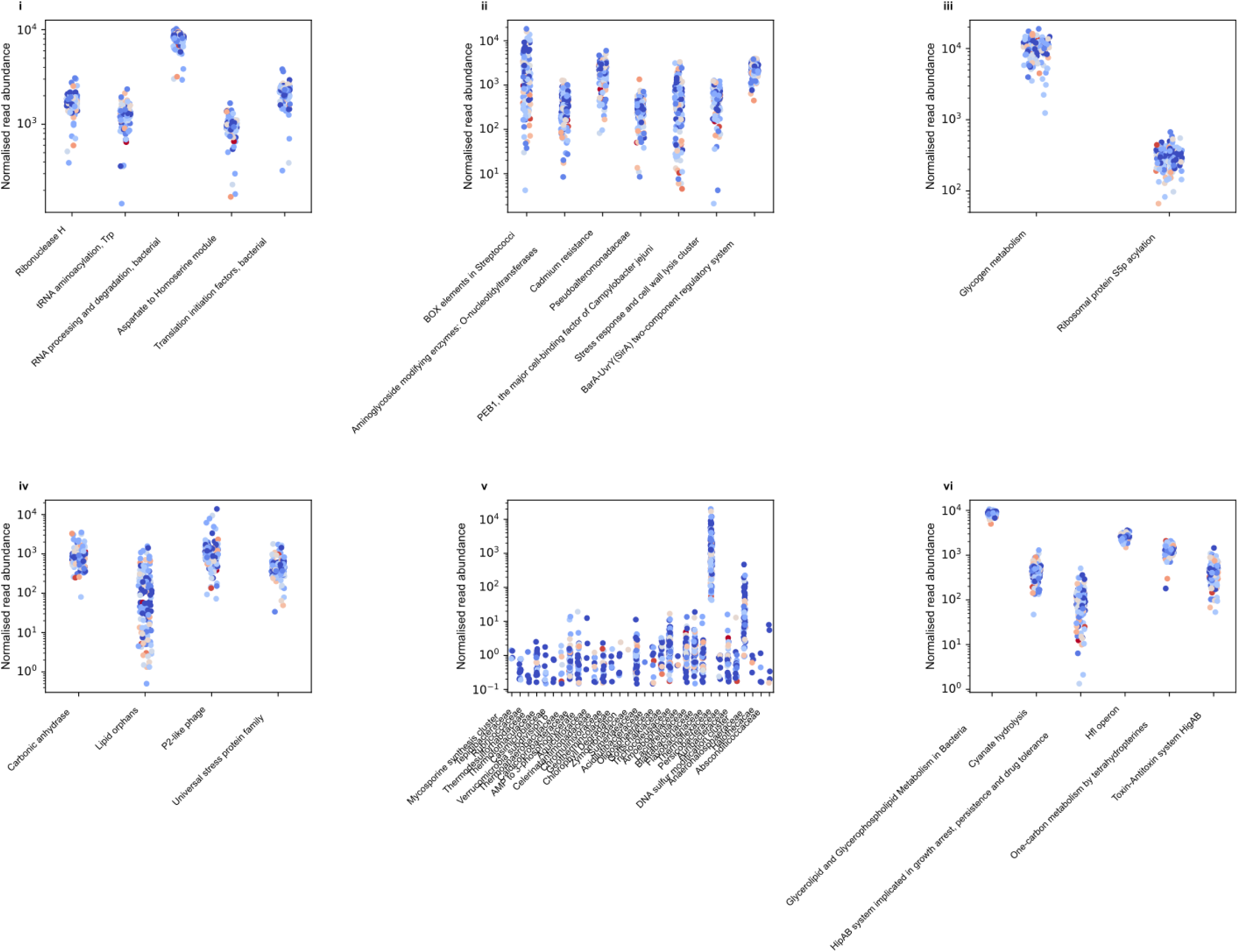
The normalised read abundance for each of the functions and phylogenies. The abundance is coloured by medication score from blue (least) to red (highest medication score). i) COPF 14; ii) COPF 6; iii) COPF 107; iv) COPF 29; v) COPF 138; vi) COPF 132 (*n* = 125 for each panel).

**Supplementary Fig. 6.**
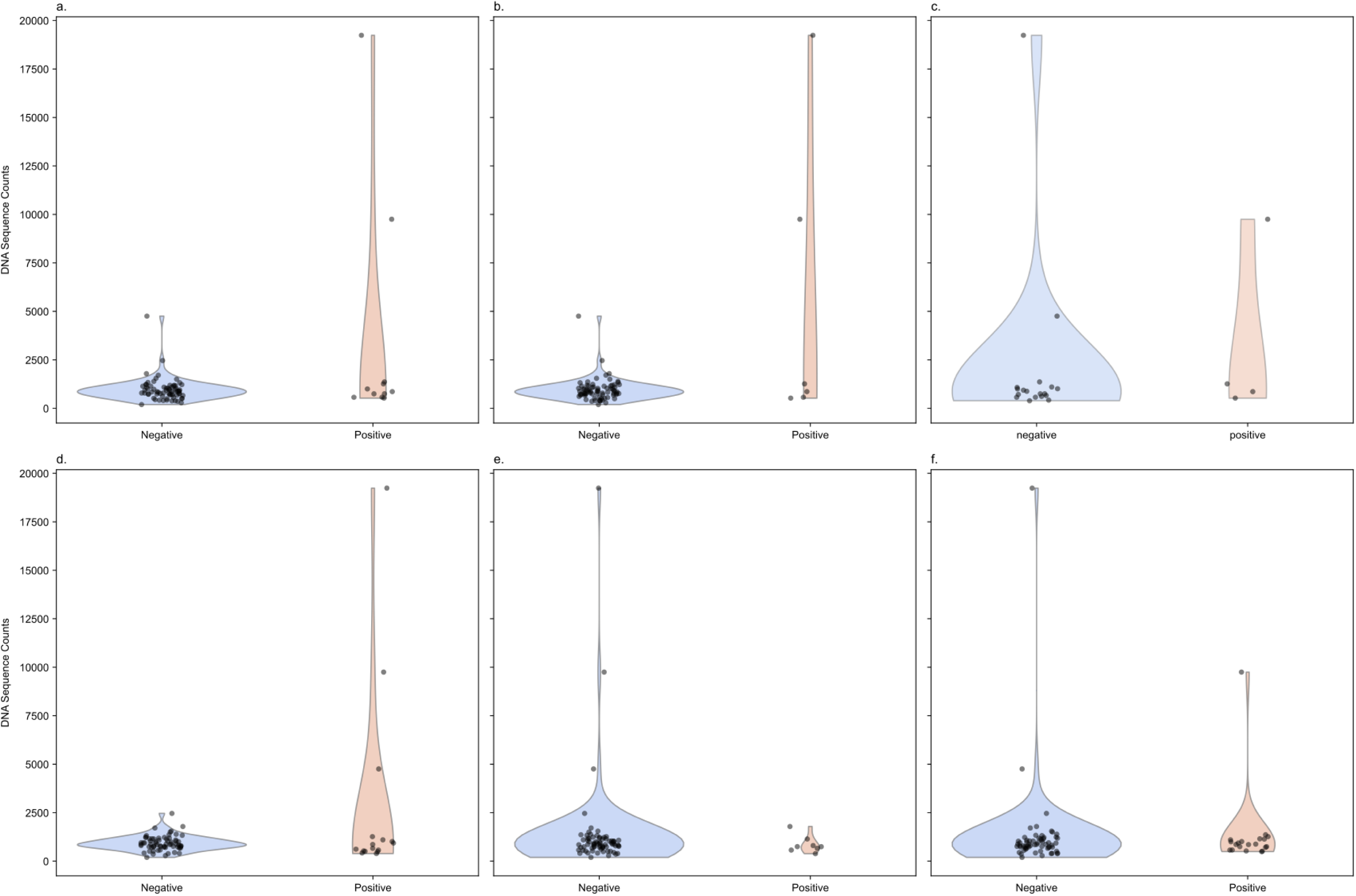
Culture of NTM compared to number of sequences detected. **a.** NTM culture status (n=82), **b**. *Mycobacteroides abscessus* culture status (n=82), **c.** NTM smear status (n=22), **d.** Rapid-growing NTM culture within the past year (n=82), **e.** Slow-growing NTM culture within the past year (n=82), **f.** Positive over the next 12 months (n=82)

**Supplementary Fig. 7.**
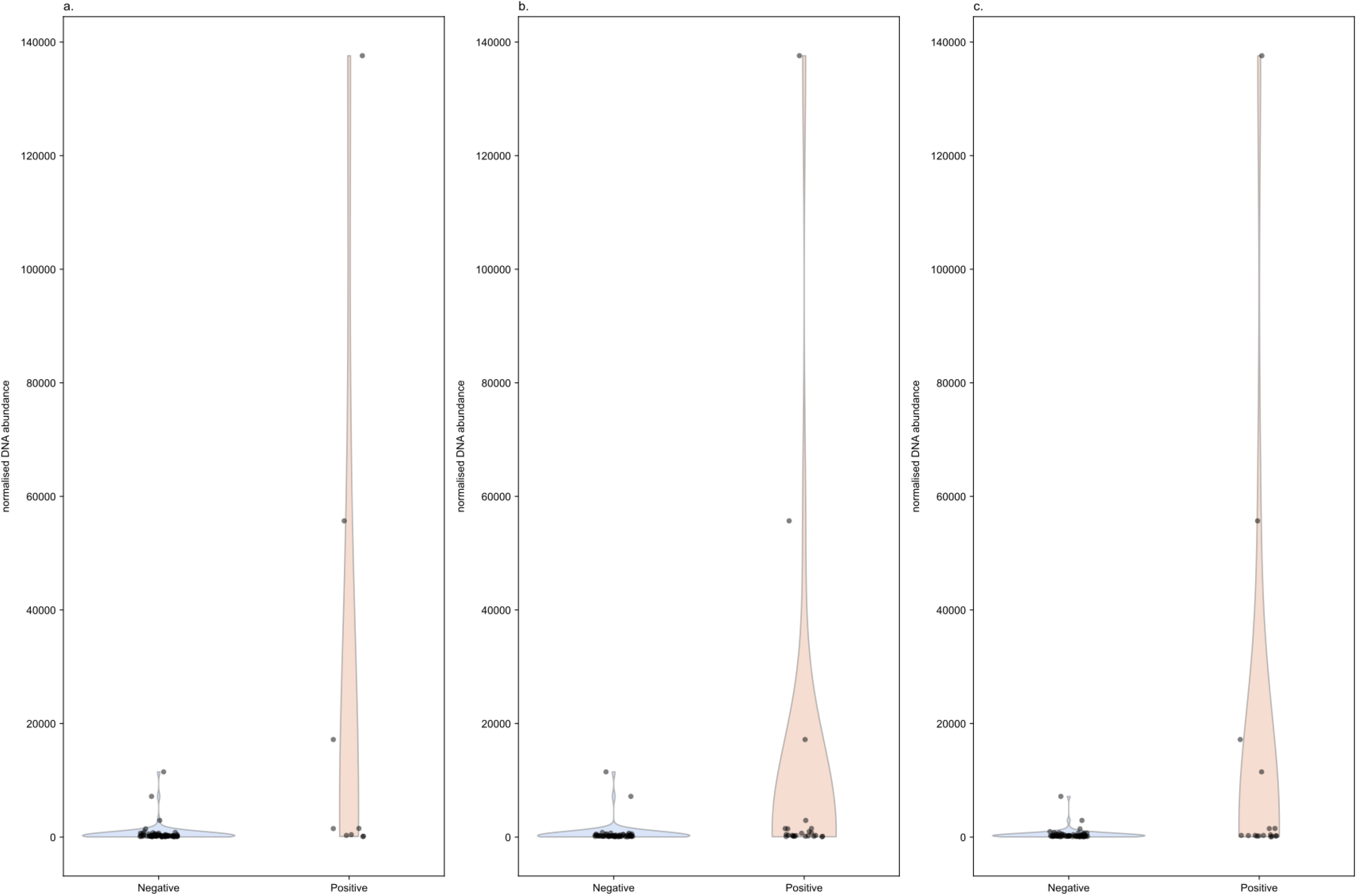
*Stenotrophomonas* culturing vs. sequence read abundance. **a.** *Stenotrophomonas maltophilia* culture status, **b.** *Stenotrophomonas maltophilia* culture status in the previous 12 months, **c.** *Stenotrophomonas maltophilia* culture status in the next 12 months (*n*=81).

**Supplementary Fig. 8.**
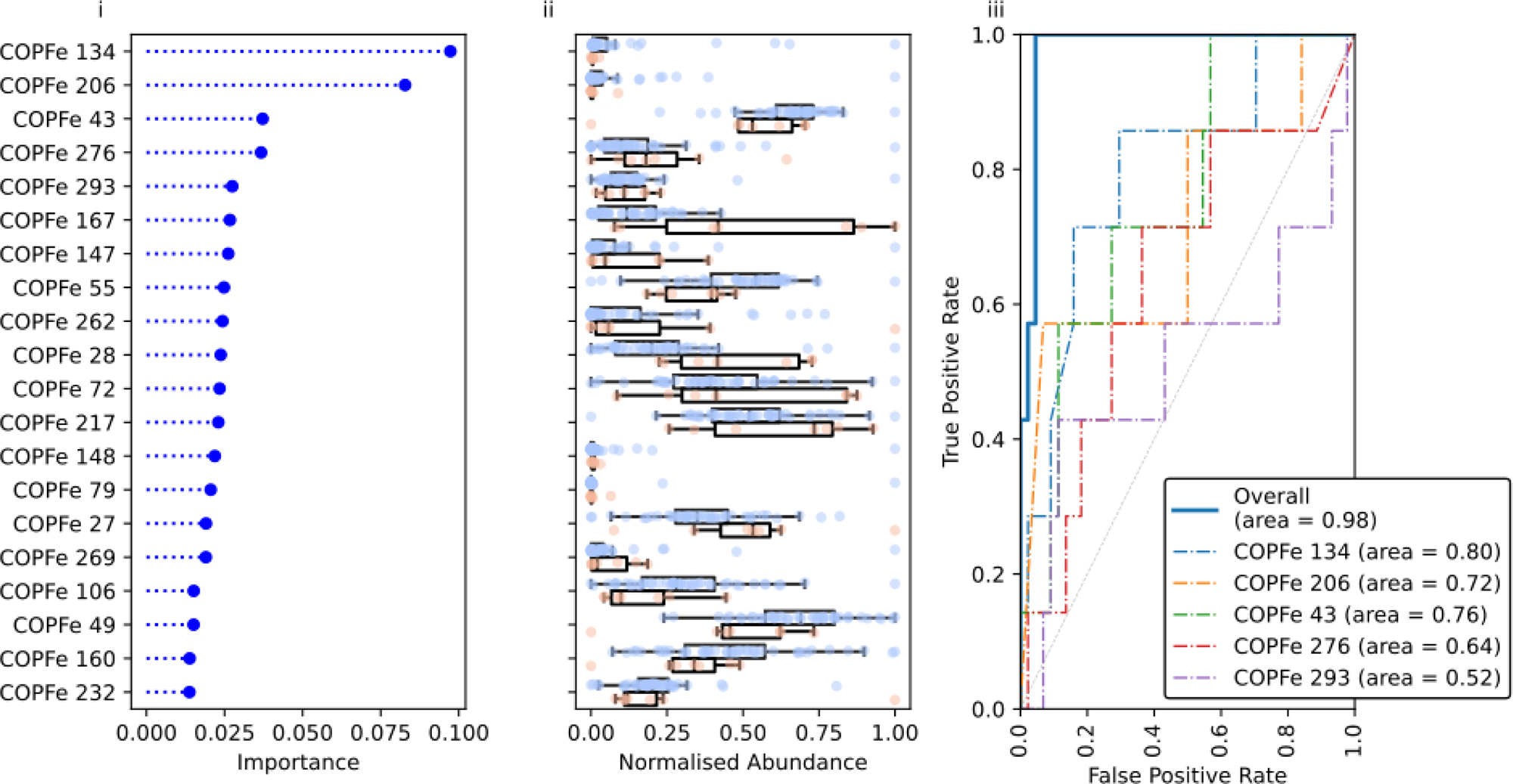
Detection of Allergic Bronchopulmonary Aspergillosis. The triptych contains (i) the gradient-boosted random forest feature importance plot for each eukaryotic COPF (called COPFe), (ii) the abundance of those COPFes in samples with (red) or without (blue) ABPA, and (iii) the ROC curve for the top 5 COPFes at predicting the phenotype and the overall ROC curve for all data. Area under the curves (AUC) are included in the ROC legend (*n* = 51).

## Notes

### Competing Interest Statement

The authors have declared no competing interest.

## References

1. Steinberg, R. et al. Longitudinal effects of elexacaftor/tezacaftor/ivacaftor on the oropharyngeal metagenome in adolescents with cystic fibrosis. J. Cyst. Fibros. 24, 562–570 (2025).

2. Martin, C. et al. Longitudinal microbial and molecular dynamics in the cystic fibrosis lung after Elexacaftor-Tezacaftor-Ivacaftor therapy. Respir. Res. 24, 317 (2023).

3. Lim, Y. W. et al. Metagenomics and metatranscriptomics: windows on CF-associated viral and microbial communities. J. Cyst. Fibros. 12, 154–164 (2013).

4. Cromwell, E. A. et al. Cystic fibrosis prevalence in the United States and participation in the Cystic Fibrosis Foundation Patient Registry in 2020. J. Cyst. Fibros. 22, 436–442 (2023).

5. UK Cystic Fibrosis Registry 2024 Annual Data Report. (2025).

6. Coburn, B. et al. Lung microbiota across age and disease stage in cystic fibrosis. Sci. Rep. 5, 10241 (2015).

7. Edwards, R. Linsalrob/Atavide_lite: Atta Pony. (Zenodo, 2025). doi:10.5281/ZENODO.15356766.

8. Robert Edwards, R. CF_Data_Analysis. (2025). doi:10.5281/zenodo.1234.

9. Zhou, S., Liu, B., Zheng, D., Chen, L. & Yang, J. VFDB 2025: an integrated resource for exploring anti-virulence compounds. Nucleic Acids Res. 53, D871–D877 (2025).

10. Lim, Y. W. et al. Clinical insights from metagenomic analysis of sputum samples from patients with cystic fibrosis. J. Clin. Microbiol. 52, 425–437 (2014).

11. Benyamini, P. Phylogenetic tracing of evolutionarily conserved Zonula occludens toxin reveals a “high value” vaccine candidate specific for treating multi-strain Pseudomonas aeruginosa infections. Toxins (Basel) 16, 271 (2024).

12. Diederich, C. et al. Mechanisms and specificity of phenazine biosynthesis protein PhzF. Sci. Rep. 7, 6272 (2017).

13. Whiteson, K. L. et al. Breath gas metabolites and bacterial metagenomes from cystic fibrosis airways indicate active pH neutral 2,3-butanedione fermentation. ISME J. 8, 1247–1258 (2014).

14. Visca, P., Imperi, F. & Lamont, I. L. Pyoverdine siderophores: from biogenesis to biosignificance. Trends Microbiol. 15, 22–30 (2007).

15. Létoffé, S. et al. Pseudomonas aeruginosa production of hydrogen cyanide leads to airborne control of Staphylococcus aureus growth in biofilm and in vivo lung environments. MBio 13, e0215422 (2022).

16. Kondakova, T. et al. Glycerophospholipid synthesis and functions in Pseudomonas. Chem. Phys. Lipids 190, 27–42 (2015).

17. Pribat, A. et al. FolX and FolM are essential for tetrahydromonapterin synthesis in Escherichia coli and Pseudomonas aeruginosa. J. Bacteriol. 192, 475–482 (2010).

18. Wood, T. L. & Wood, T. K. The HigB/HigA toxin/antitoxin system of Pseudomonas aeruginosa influences the virulence factors pyochelin, pyocyanin, and biofilm formation. Microbiologyopen 5, 499–511 (2016).

19. Klockgether, J., Cramer, N., Wiehlmann, L., Davenport, C. F. & Tümmler, B. Pseudomonas aeruginosa Genomic Structure and Diversity. Front. Microbiol. 2, 150 (2011).

20. Hreha, T. N. et al. The three NADH dehydrogenases of Pseudomonas aeruginosa: Their roles in energy metabolism and links to virulence. PLoS One 16, e0244142 (2021).

21. Santi, I., Manfredi, P., Maffei, E., Egli, A. & Jenal, U. Evolution of antibiotic tolerance shapes resistance development in chronic Pseudomonas aeruginosa infections. MBio 12, (2021).

22. Marvig, R. L., Sommer, L. M., Molin, S. & Johansen, H. K. Convergent evolution and adaptation of Pseudomonas aeruginosa within patients with cystic fibrosis. Nat. Genet. 47, 57–64 (2015).

23. Daley, C. L. et al. Treatment of non-tuberculous Mycobacterial pulmonary disease: An official ATS/ERS/ESCMID/IDSA clinical practice guideline. Clin. Infect. Dis. 71, e1–e36 (2020).

24. Agarwal, R. et al. Revised ISHAM-ABPA working group clinical practice guidelines for diagnosing, classifying and treating allergic bronchopulmonary aspergillosis/mycoses. Eur. Respir. J. 63, 2400061 (2024).

25. Feigelman, R. et al. Sputum DNA sequencing in cystic fibrosis: non-invasive access to the lung microbiome and to pathogen details. Microbiome 5, 20 (2017).

26. Nelson, M. T. et al. Human and Extracellular DNA Depletion for Metagenomic Analysis of Complex Clinical Infection Samples Yields Optimised Viable Microbiome Profiles. Cell Rep. 26, 2227–2240.e5 (2019).

27. Dmitrijeva, M. et al. Strain-Resolved Dynamics of the Lung Microbiome in Patients with Cystic Fibrosis. MBio 12, (2021).

28. Motta, H. et al. Comparative microbiome analysis in cystic fibrosis and non-cystic fibrosis bronchiectasis. Respir. Res. 25, 211 (2024).

29. Hilliam, Y. et al. Cystic fibrosis pathogens persist in the upper respiratory tract following initiation of elexacaftor/tezacaftor/ivacaftor therapy. Microbiol. Spectr. 12, e0078724 (2024).

30. Armbruster, C. R. et al. Persistence and evolution of Pseudomonas aeruginosa following initiation of highly effective modulator therapy in cystic fibrosis. MBio 15, e0051924 (2024).

31. Mostacci, N. et al. Informed interpretation of metagenomic data by StrainPhlAn enables strain retention analyses of the upper airway microbiome. mSystems e0072423 (2023).

32. Pallenberg, S. T. et al. Impact of Elexacaftor/Tezacaftor/Ivacaftor Therapy on the Cystic Fibrosis Airway Microbial Metagenome. Microbiol Spectr 10, e0145422 (2022).

33. Van den Bossche, S., De Broe, E., Coenye, T., Van Braeckel, E. & Crabbé, A. The cystic fibrosis lung microenvironment alters antibiotic activity: causes and effects. Eur. Respir. Rev. 30, 210055 (2021).

34. Crabbé, A., Jensen, P. Ø., Bjarnsholt, T. & Coenye, T. Antimicrobial tolerance and metabolic adaptations in microbial biofilms. Trends Microbiol. 27, 850–863 (2019).

35. Flinders University. Deep Thought (HPC). Preprint at 10.25957/FLINDERS.HPC.DEEPTHOUGHT (2021).

36. Pawsey Supercomputing Research Centre. Setonix. Preprint at 10.48569/18SB-8S43 (2023).

37. Chen, S., Zhou, Y., Chen, Y. & Gu, J. fastp: an ultra-fast all-in-one FASTQ preprocessor. Bioinformatics 34, i884–i890 (2018).

38. Li, H. Minimap2: pairwise alignment for nucleotide sequences. Bioinformatics 34, 3094–3100 (2018).

39. Li, H. et al. The Sequence Alignment/Map format and SAMtools. Bioinformatics 25, 2078–2079 (2009).

40. Suzek, B. E. et al. UniRef clusters: a comprehensive and scalable alternative for improving sequence similarity searches. Bioinformatics 31, 926–932 (2015).

41. Shen, W. & Ren, H. TaxonKit: A practical and efficient NCBI taxonomy toolkit. J. Genet. Genomics 48, 844–850 (2021).

42. Olson, R. D. et al. Introducing the Bacterial and Viral Bioinformatics Resource Center (BV-BRC): a resource combining PATRIC, IRD and ViPR. Nucleic Acids Res. 51, D678–D689 (2023).

43. Li, D., Liu, C.-M., Luo, R., Sadakane, K. & Lam, T.-W. MEGAHIT: an ultra-fast single-node solution for large and complex metagenomics assembly via succinct de Bruijn graph. Bioinformatics 31, 1674–1676 (2015).

44. Nissen, J. N. et al. Improved metagenome binning and assembly using deep variational autoencoders. Nat. Biotechnol. 39, 555–560 (2021).

45. Kang, D. D. et al. MetaBAT 2: an adaptive binning algorithm for robust and efficient genome reconstruction from metagenome assemblies. PeerJ 7, e7359 (2019).

46. Alneberg, J. et al. Binning metagenomic contigs by coverage and composition. Nat. Methods 11, 1144–1146 (2014).

47. Roach, M. J. et al. Koverage: Read-coverage analysis for massive (meta)genomics datasets. J. Open Source Softw. 9, 6235 (2024).

48. Pedregosa, F. et al. Scikit-learn: Machine Learning in Python. J. Mach. Learn. Res. abs/1201.0490, 2825–2830 (2011).

49. Abadi, M. et al. TensorFlow, Large-Scale Machine Learning on Heterogeneous Systems. (2015). doi:10.5281/zenodo.4724125.

