## Supplemental Tables and Information for "Microbial Ecological Signatures Predict Pathogen Emergence and Multidrug Resistance in Cystic Fibrosis Airways up to a Year in Advance"

#### Supplemental Online Material

### Supplemental Results Tables

**Table S1.** Number of entries in the functional (subsystems) data at different levels and number of metagenome assembled genomes. We used the subsystems level for most analyses.

| Hierarchical level | Number of entries |
|---|---|
| **Classes** | 11 |
| **Level 1** | 29 |
| **Level 2** | 139 |
| **Subsystems** | 769 |
| **All functions** | 6,229 |
| **Metagenome Assembled Genomes (MAGs)** | 3,214 |

**Table S2.** Number of entries at different taxonomic levels for either just the Bacteria or all taxa (including Archaea, Bacteria, Eukarya, and Viruses). We used the Family level taxonomy for most analyses.

| Taxonomy | Bacteria | All taxa |
|---|---|---|
| **Kingdom** | 1 | 4 |
| **Phylum** | 164 | 252 |
| **Class** | 140 | 348 |
| **Order** | 302 | 837 |
| **Family** | 743 | 1,857 |
| **Genus** | 3,581 | 5,806 |
| **Species** | 26,499 | 31,371 |

**Table S3.** Features (taxonomy and subsystems) present in COPF 31, the most predictive cluster for *Pseudomonas* colonisation.

| | | |
|---|---|---|
| ABC-type iron transport system | Geothermobacteraceae | Potential DNA repair cluster |
| Acyclic terpene utilisation | Heme and heme d1 biosynthesis from siroheme | Pseudomonadaceae |
| Adenylylsulfate reductase | IbrA and IbrB: co-activators of prophage gene expression | Respiratory Systems - Shewanella (updated) |
| Aestuariirhabdaceae | L-2-amino-4-methoxy-trans-3-butenoic acid synthesis | Rhamnolipids in Pseudomonas |

| | | |
|---|---|---|
| Beta-lactamases Ambler class D | Malonate decarboxylase | Salmochelin-mediated Iron Acquisition |
| Biogenesis of cbb3-type cytochrome c oxidases | Marinobacteraceae | Siderophore Pyoverdine |
| Cholera toxin | MexXY System of Pseudomonas aeruginosa | Stress proteins YciF, YciE |
| CMP-pseudaminic acid synthesis | Mycobacterial FadD proteins (fatty acid CoA- and AMP- ligases) | Sulfate assimilation related cluster |
| Coenzyme PQQ synthesis | Mycobacterial FadE proteins Acyl-CoA dehydrogenase | Sulfur transfer pathway CsdAEL |
| Colwelliaceae | Mycobacterial gene cluster associated with resistance against FAS-II antibiotics | Trans-envelope signaling system VreARI in Pseudomonas |
| Cytochrome c oxidase EC 1.9.3.1 | Outer membrane porins in Pseudomonas and Acinetobacter | Two partner secretion pathway (TPS) |
| Extracellular matrix proteins (PEL) involved in glucose-rich biofilm formation in Pseudomonas | Periplasmic nitrate reductase EC 1.7.99.4 | Type 4 conjugative transfer system, IncI1 type |
| Extracellular matrix proteins (PSL) involved in mannose-rich biofilm formation in Pseudomonas | Phenazine biosynthesis | Type III secretion system |

**Table S4.** Features (taxonomy and subsystems) present in COPF 132, a predictive cluster for *Pseudomonas* colonisation.

| | | |
|---|---|---|
| Glycerolipid and Glycerophospholipid Metabolism in Bacteria | Cyanate hydrolysis | HipAB system implicated in growth arrest, persistence and drug tolerance |
| Hfl operon | One-carbon metabolism by tetrahydropterines | Toxin-Antitoxin system HigAB |

**Table S5.** Features important for predicting MDR *Pseudomonas*

| COPF | Features | Higher in |
|---|---|---|
| **29** | Carbonic anhydrase; Lipid orphans; P2-like phage, Universal stress protein family | Non-MDR |
| **68** | DNA processing cluster, Calvin-Benson cycle, Peptidase clustering with DAP, De Novo Pyrimidine Synthesis. | MDR |
| **82** | NADH ubiquinone oxidoreductase, Menaquinone biosynthesis from chorismate via 1,4-dihydroxy-6-naphthoate | Non-MDR |
| **18** | Fe-S cluster assembly, Streptococcal Hyaluronic Acid Capsule, Aspartate to Threonine Module, rRNA modification related cluster including tlyA, Translation termination factors, bacterial, RNA polymerase, bacterial, Sulfoquinovose biosynthesis cluster. | MDR |

**Table S6.** Features important for predicting conversion to *Pseudomonas* culture positivity in the next 12 months.

| COPF | Features | Higher in |
|---|---|---|
| **110** | Cell wall-associated cluster in Mycobacterium, 'Xanthine dehydrogenase subunits, Iron(III) dicitrate transport system Fec, Gemmatimonadaceae | |
| **24** | Phenylalanine and Tyrosine synthesis 1, DeNovo Purine Biosynthesis, Lysine DAP biosynthetic pathway, Diaminopimelate Synthesis, Lysine leader peptide | |
| **11** | Enoyl-[ACP] reductases disambiguation, NAD and NADP cofactor biosynthesis global, NAD and NADP cofactor biosynthesis bacterial, Fatty Acid Biosynthesis cluster, Resistance to Triclosan | |

**Table S7.** Number of culture positive samples out of 127 samples in total

| Culture Status | Number of positive samples |
|---|---|
| *Achromobacter xylosoxidans* | 3 |
| *Acremonium* species | 1 |
| *Aspergillus flavus* | 3 |
| *Aspergillus fumigatus* | 22 |
| *Aspergillus nidulans* | 2 |
| *Aspergillus niger* | 1 |
| *Aspergillus terreus* | 1 |
| *Burkholderia cepacia* | 1 |
| *Candida albicans* | 40 |
| *Haemophilus influenzae* | 1 |
| *Inquilinus limosus* | 1 |
| *Lomentospora prolificans* | 1 |
| *Oral flora* | 31 |
| *Penicillium* | 3 |
| *Scedosporium apiospermum* | 1 |
| *Staphylococcus aureus* | 28 |
| *Stenotrophomonas maltophilia* | 9 |
| **Pseudomonas** | |
| *Pseudomonas aeruginosa* | 36 |
| MDR *Pseudomonas aeruginosa* | 14 |
| mucoid | 19 |
| non-mucoid | 13 |

**Table S8.** *Mycobacteria* observations in our cohort

| pwCF Identifier | Days Since First Sample | Smear status | Cultured *Mycobacteria* |
|---|---|---|---|
| 642660 | 256 | negative | *M. abscessus; M. intracellulare* |
| 670829 | 13 | positive | *M. abscessus* |
| 698564 | 443 | negative | Unclassified rapid NTM (smear negative) - previously identified as *M. abscessus* (smear negative) |
| 698917 | 80 | negative | *M. abscessus* |
| 717449 | 261 | positive | *M. abscessus* |
| 720054 | 7 | negative | *M. intracellulare* |
| 720054 | 63 | negative | *M. intracellulare* |
| 753522 | 261 | positive | *M. abscessus* |
| 763742 | 7 | negative | unspecified NTM |
| 770560 | 449 | positive | *M. abscessus* |
| 1162967 | 120 | negative | *M. terrae* (intermediate grower) |

**Table S9.** The *Aspergillus* culture status and sequence detection of pwCFID 676138 over almost 200 days.

| | | ***Metagenome sequence abundances***[1] | | | | | | | ***Culture Status*** | | | | | ***Aspergillus fumigatus culturing*** | |
|---|---|---|---|---|---|---|---|---|---|---|---|---|---|---|---|
| **Days Since First Sample** | **Sum of antifungals** | ***A. candidus*** | ***A. fischeri*** | ***A. fumigatus*** | ***A. mulundensis*** | ***A. sclerotialis*** | ***A. turcosus*** | ***A. udagawae*** | ***A. fumigatus*** | ***A. flavus*** | ***A. nidulans*** | ***A. niger*** | ***A. terreus*** | **Previous 12 months** | **Next 12 months** |
| 13 | 0 | 0.0 | 0.0 | 94.95 | 0.0 | 0.0 | 0.0 | 1.76 | - | - | - | - | - | Yes | Yes |
| 21 | 0 | 0.0 | 0.41 | 2.86 | 0.0 | 0.0 | 0.0 | 0.0 | Yes | - | - | - | - | Yes | Yes |
| 134 | 2 | 0.0 | 0.0 | 18.37 | 0.28 | 0.0 | 0.0 | 0.0 | - | - | - | - | - | Yes | Yes |
| 189 | 1 | 0.0 | 0.0 | 30.44 | 0.0 | 0.0 | 0.0 | 0.0 | Yes | - | - | - | - | Yes | Yes |
| 199 | 2 | 0.17 | 0.0 | 1.74 | 0.0 | 0.17 | 0.52 | 0.0 | - | - | - | - | - | Yes | Yes |

[1]Normalised abundance of each taxa identified from the short-read sequences.

**Table S10.** Allergic Bronchopulmonary Aspergillosis status for our cohort

| ABPA status | Samples | Unique pwCF |
|---|---|---|
| **Not sensitised** | 18 | 15 |
| **Sensitised** | 26 | 18 |
| **ABPA** | 7 | 4 |

**Table S11.** Accuracy of predicting *Pseudomonas* culturability in worldwide samples.

| Project Title | Sample | Runs Analysed | Runs Correctly Predicted | True Positive Predictions (TP) | False Negative Preductions (FN) | True Negative Predictions (TN) | False Positive Predictions (FP) |
|---|---|---|---|---|---|---|---|
| Sputum Metagenome Of CF, COPD, Smokers And Healthy Subjects | PRJNA316588 | 17 | 17 | 2 | 0 | 15 | 0 |
| Pseudomonas aeruginosa. Sinus, throat, and sputum airway microbiology in cystic fibrosis | PRJNA1081394 | 330 | 311 | 310 | 19 | 1 | 0 |
| Depletion of human DNA from complex clinical samples for metagenomic sequencing | PRJNA516442 | 15 | 15 | 8 | 0 | 7 | 0 |
| Shotgun metagenomic data from bronchoalveolar lavage fluid (BALF) samples of infants with CF | PRJNA1126024 | 2 | 2 | 1 | 0 | 1 | 0 |
| Impact of ELX/TEZ/IVA therapy on the CF airway metagenome | PRJEB51171 | 9 | 8 | 3 | 1 | 5 | 0 |
| Time series of patients with cystic fibrosis | PRJNA510441 | 14 | 5 | 5 | 9 | 0 | 0 |
| *Pseusomonas aeruginosa* metagenomes | PRJEB14440 | 5 | 5 | 5 | 0 | 0 | 0 |
| Microbiome analysis in the respiratory tract of individuals with cystic fibrosis and non-cystic fibrosis bronchiectasis | PRJNA1055940 | 61 | 45 | 9 | 16 | 36 | 0 |
| Lung Microbiome Dynamics in Cystic Fibrosis | PRJEB32062 | 25 | 24 | 16 | 1 | 8 | 0 |
| Study of fungal clinical isolates from different anatomical sites and metagenomics of the lung microbiome of patients with cystic fibrosis | PRJNA644285 | 12 | 11 | 3 | 1 | 8 | 0 |
| The metagenome of children with cystic fibrosis | PRJNA931830 | 260 | 259 | 11 | 0 | 248 | 1 |

| | | | | | | |
|---|---|---|---|---|---|---|
| **Total: Confident samples** | **750** | **702** | **373** | **47** | **329** | **1** |

**Table S12.** Accuracy of predicting *Pseudomonas* culturability in worldwide samples with uncertain provenance.

| Project Title | Sample | Runs Analysed | Runs Correctly Predicted (TP) | Comments |
|---|---|---|---|---|
| Longitudinal changes on the oropharyngeal metagenome among children with cystic fibrosis after ETI | PRJNA1101448 | 323 | - | It is not clear how many have Pseudomonas |
| Cystic fibrosis pathogens | PRJEB20836 | 8 | 8 | |
| Viral and Microbial Cystic Fibrosis Lung Metagenome | PRJNA71831 | 21 | 4 | It is not clear which samples, if any, have Pseudomonas |
| Sputum Metagenomics | PRJEB54014 | 75 | 21 | There are no culture reports described in the paper |
| Cystic fibrosis airway respiratory microbiome | PRJNA615628 | 71 | 2 | They culture Pseudomonas from 8 samples but do not specify which samples. |
| CF airway microbiome | PRJNA825831 | 114 | 12 | In the paper they report 47% Pseudomonas positivity, but also describe 20/22 pwCF with Pseudomonas |
| Longitudinal Cystic Fibrosis Airway Raw sequence reads | PRJNA846291 | 98 | 1 | No comment in the paper on positivity |
| Cystic Fibrosis Lung Sputum Microbiome before and after antibiotic treatment | PRJNA839435 | 12 | 11 | There is no connection between SRA IDs and the IDs in the paper |
| Time-resolved study on cystic fibrosis airways | PRJNA516870 | 67 | 27 | Unpublished |
| Metagenomic data from sputa from cystic fibrosis patients | PRJNA1091195 | 22 | 12 | Unpublished |
| Sputum samples from cystic fibrosis patients' metagenomes | PRJNA316056 | 12 | 6 | Unpublished |
| **Total: Samples with uncertain provenance** | | **824** | **104** | |

#### Comparison of Sequencing Technologies

We compared the taxa identified using atavide with the different sequencing technologies (Fig. S1). The companion notebook is available as a Jupyter notebook at MGI-MinION-Promethion_Compared.ipynb. We only show the data from the ten samples that were sequenced using all three technologies (x-axis). The normalised taxa from the MGI read abundance is shown on the y-axis. We filtered the data so that only those taxa with at least 10 reads per million mapped reads are shown, which omits the long tail of organisms that have very few reads mapped and that maybe of spurious value.

### Supplemental Methods

#### Gradient Boosted Random Forest

Gradient-boosted random forests were trained with early stopping to avoid overfitting, with training halted when no further improvement in validation loss was observed.

**Table S13**. Gradient boosted random forest metrics.

| Prediction | # Negative samples | # Positive samples | Early Stopping | Model MSE† |
|---|---|---|---|---|
| ***Pseudomonas* culture positivity** | 91 | 36 | 349 | 0.19 |
| **MDR Pseudomonas** | 22 | 14 | 100 | 0.375 |
| **Conversion to *Pseudomonas* culture positivity** | 38 | 36 | 11 | 0.67 |
| **Mucoid *Pseudomonas* status** | 10 | 16 | 100 | 0.5 |

†Mean squared error

The mean squared error (MSE) for our gradient-boosted models varied widely and are reported in the table above. The MSE provides a measure of residual variance: values closer to zero indicate tighter predictions around the true outcome, whereas higher values suggest weaker fit. In our approach, these differences largely reflect the type of outcome being modelled and the degree of biological signal present in the latent clusters. Predictions of strongly constrained outcomes, such as specific antibiotic exposures, yielded very low MSEs (0.01–0.23), whereas more diffuse or heterogeneous phenotypes (e.g. complex clinical states) naturally produced higher errors.

Higher MSEs indicate that the phenotype is only partially explained by the microbiome features captured here. Given the modest cohort size, the high dimensionality of the input space, and the intrinsic stochasticity of microbial communities, this variability is expected. However, the signal is consistent across repeated runs, feature selection using the autoencoder latent dimensions is stable, and the ROC curves show discrimination above chance. Instead, the MSE reflects the biological heterogeneity of the outcomes being modelling. Strongly predictive traits give tight errors; diffuse traits produce looser fits.
